# Association between patient race/ethnicity and invasive ventilation in hypoxemic respiratory failure

**DOI:** 10.1101/2022.10.31.22281761

**Authors:** Fred M. Abdelmalek, Federico Angriman, Julie Moore, Kuan Liu, Lisa Burry, Laleh Seyyed-Kalantari, Sangeeta Mehta, Judy Gichoya, Leo Anthony Celi, George Tomlinson, Michael Fralick, Christopher J. Yarnell

## Abstract

**Importance:** Differential use of therapies for respiratory failure according to patient race/ethnicity may represent health inequity and could impact patient survival.

**Objective:** Measure the association between patient race/ethnicity and the use of invasive ventilation, and the impact of any association on survival.

**Design:** Retrospective cohort analysis using a Bayesian multistate model that adjusted for baseline covariates and time-varying severity.

**Setting:** Multicenter study using the Medical Information Mart for Intensive Care IV (MIMIC-IV) and Phillips eICU (eICU) databases from the USA.

**Participants:** Non-intubated adults receiving oxygen within the first 24 hours of ICU admission.

**Exposure:** Patient race/ethnicity (Asian, Black, Hispanic, white).

**Main outcomes and measures:** Primary output was the cause-specific hazard ratio (HR) of invasive ventilation for patient race/ethnicity. Secondary output was change in 28-day survival mediated by differences in invasive ventilation rate. We reported posterior means and 95% credible intervals (CrI).

**Results:** We studied 38,263 patients, 52% (20,033) from MIMIC-IV and 48% (18,230) from eICU, 2% Asian (892), 11% Black (4,289), 5% Hispanic (1,964), and 81% white (31,923). Invasive ventilation occurred in 3,511 (9.2%), and 2,869 (7.5%) died. The rate of invasive ventilation was lower in Asian (HR 0.82, CrI 0.70 to 0.95), Black (HR 0.78, CrI 0.71 to 0.86), and Hispanic (HR 0.70, CrI 0.61 to 0.79) patients as compared to white patients. For the average patient, lower rates of invasive ventilation did not mediate differences in survival. For a reference patient with inspired oxygen (FiO2) varied from 0.5 to 1.0, the change in survival mediated by lower rates of invasive ventilation ranged from probable benefit (probability 0.82 for Asian patients, 0.91 for Black patients, and 0.93 for Hispanic patients) at FiO2 0.5 to probable harm (probability 0.87 for Asian patients, 0.92 for Black patients, and 0.97 for Hispanic patients) at FiO2 1.0, although the mean absolute changes in mortality were all less than 1.5%.

**Conclusions:** Asian, Black, and Hispanic patients had a lower rate of invasive ventilation than white patients. The changes in 28-day survival mediated by this difference ranged from slight benefit at lower inspired oxygen fractions to slight harm at inspired oxygen fraction of 1.0, and there was no difference in survival for the average patient.

**Key Points:** *Question:* What is the association between patient race/ethnicity and the use of invasive ventilation in hypoxemic respiratory failure, and what is the impact of any differences on survival?

*Findings:* We studied 38,263 patients from two US databases, who were 2% Asian (892), 11% Black (4,289), 5% Hispanic (1,964), and 81% white (31,118). Invasive ventilation occurred in 3,511 (9.2%), and 2,869 (7.5%) died. The hazard ratio (HR) for invasive ventilation was lower in Asian (HR 0.82, CrI 0.70 to 0.95), Black (HR 0.78, CrI 0.71 to 0.86), and Hispanic (HR 0.70, CrI 0.61 to 0.79) patients as compared to white patients. For the average patient, race/ethnicity differences in invasive ventilation rates did not mediate differences in 28-day survival. For the reference patient, at inspired oxygen fractions up to 0.9, lower invasive ventilation rates mediated a modest survival benefit, whereas at inspired oxygen fraction of 1.0, the lower invasive ventilation rates mediated a modest survival decrease, although the absolute changes were all less than 1.5%.

*Meaning:* Asian, Black, and Hispanic patients had a lower rate of invasive ventilation than white patients. Although this difference had no impact on 28-day survival for the average patient, the change in survival mediated by lower rates of invasive ventilation could range from slight benefit at lower inspired oxygen fractions to slight harm at inspired oxygen fraction of 1.0.

## Introduction

Outcomes for patients with acute respiratory failure vary according to patient race/ethnicity. Across racial/ethnic categories, age-adjusted respiratory failure-related mortality in the United States is highest among non-Hispanic Black patients.^1^ Black and Hispanic patients hospitalized with respiratory failure face a higher odds of in-hospital death than White patients.^2^ Among patients enrolled in clinical trials, Black or Hispanic race/ethnicity as opposed to White race/ethnicity is associated with higher mortality.^3^ Asian patients admitted to intensive care units (ICUs) also have higher age- and sex-adjusted mortality rates.^4^

Mechanisms for these disparities are not fully known.^5^ Peripheral oximeters can overestimate arterial oxygen saturation in patients with darker skin pigment, leading to delayed initiation of supplemental oxygen.^6–9^ Beginning in childhood, Black and Hispanic children have disproportionate exposure to air pollution.^10^ Differences in care preferences may contribute.^11^ Decreased access to primary and preventative care among Black patients may lead to more critical illness.^12^ In the USA, Black and Hispanic individuals are disproportionately cared for in minority-serving hospitals, where mortality has shown less improvement over time compared to mortality in non-minority-serving hospitals.^13^ Last, implicit bias is known to influence clinical decision-making.^14^

Differences in rates of invasive ventilation may partially explain observed disparities in mortality. Invasive ventilation is a potentially life-saving intervention for patients with severe respiratory failure, but there are no consensus criteria for its use.^15–17^ The decision is influenced by hospital culture and vulnerable to implicit bias.^14^ Prior studies offer conflicting findings on the association between race/ethnicity and invasive ventilation. These studies are limited because none adjusted for time-varying severity, many used decedent cohorts, and none attempted to estimate the impact of differences on outcomes.^18–27^ We performed a retrospective cohort study with adjustment for baseline confounding and time-varying severity to investigate (1) the association between patient race/ethnicity and use of invasive ventilation and (2) the survival effect mediated by differential use of invasive ventilation.

## Methods

### Study setting and patients

We performed a retrospective cohort study of adult patients requiring supplemental oxygen within their first 24 hours of ICU admission. Patients were excluded if they had race/ethnicity classified as “Other” or “unknown”, had received invasive ventilation previously during the same ICU admission, or if they were admitted to ICU from the operating room. We combined data from two deidentified databases of patient-level clinical data: Medical Information Mart for Intensive Care version IV (MIMIC-IV) and Phillips eICU database (eICU).^28–32^ MIMIC-IV includes 76,540 ICU admissions (2008-2019) from a single academic quaternary center in Boston, USA, while eICU includes 200,859 admissions (2014-2015) from 208 hospitals across the USA participating in Phillips telemonitoring service for intensive care.

This study followed guidelines for reporting of race/ethnicity and observational study conduct.^33,34^ Use of the MIMIC-IV database for research has been approved by institutional review board of Massachusetts Institute of Technology (number 0403000206). The eICU database has been deemed to meet safe harbour standards by an independent privacy expert (Privacert, Cambridge, MA, USA; Health Insurance Portability and Accountability Act Certification number 1031219–2).^28,31^

### Variables

The exposure was patient race/ethnicity, coded as Asian, Black, Hispanic, or white. This information was entered by an administrator who asked patients or their family for their race/ethnicity.^35^ Baseline covariates were chosen to include confounders of the relationship between race/ethnicity and invasive ventilation, race/ethnicity and survival, and invasive ventilation and survival: age, sex, congestive heart failure (CHF), chronic obstructive pulmonary disease (COPD), cancer, dementia, year of ICU admission, database (MIMIC-IV or eICU), hospital region, teaching hospital status, and ICU type (cardiac, medical-surgical, or neuro-trauma).

To account for possible differences in time-varying severity by patient race/ethnicity, we included time-varying covariates: oxygen device [none, nasal prongs, facemask, non-rebreather mask, high-flow nasal cannula (HFNC), non-invasive positive pressure ventilation (NIV)], heart rate, respiratory rate, peripheral oxygen saturation, inspired oxygen fraction (FiO2), Glasgow Coma Scale (GCS), and vasopressor use. Work of breathing (normal or abnormal) and insurance information (Other, Medicaid, Medicare) were only available in MIMIC-IV data and included in a sensitivity analysis. All numerical variables were transformed, centered, and scaled (Supplement).

### Outcomes and follow-up

Invasive ventilation was the primary outcome. Hospital survival at 28 days was the secondary outcome. Patients were followed from moment of first recorded oxygen use in ICU until the earliest of death or 28 days. Patients discharged alive from hospital were assumed to have survived to 28 days.

### Analysis

The analysis was a continuous-time Bayesian multistate survival model with parametric baseline hazards. Patient states included oxygen therapy, invasive ventilation, ICU discharge, and death (Figure S1). Transitions originating from oxygen therapy used an exponential hazard function and adjustment for time-varying severity. Transitions originating from invasive ventilation or ICU discharge used a Weibull hazard function and adjustment for time of state entry. All transitions incorporated baseline covariates. We used skeptical prior distributions (normal with mean 0 and standard deviation 0.3) on a log-hazard scale for intercept and all coefficients.^36^ This expressed prior belief that a large association between any single coefficient and outcome was unlikely, and biased results away from spurious extreme associations.^36^

The primary output was the cause-specific hazard ratio for invasive ventilation according to patient race/ethnicity.^37^ The secondary output was the absolute change in survival mediated by differences in invasive ventilation rates, also known as the indirect effect. These were generated by Monte Carlo integration using Bayesian multistate modeling.^38^ The indirect effect quantifies predicted change in patient survival if non-white patients had similar probability of invasive ventilation as white patients, after adjusting for baseline covariates and time-varying severity.^38^

We reported the mediation analysis for the average patient from each race/ethnicity subgroup. Covariates for the average patients were defined by the average values of each covariate across that subgroup. We allowed for binary covariates to take the value of their proportion (for example, if 21% of patients had COPD, the COPD covariate value was set to 0.21, not 0 or 1). We also reported mediation for the average patient by race/ethnicity within subgroups of sex (male, female) and age (< 50 years, 50-65 years, 65-80 years, ≥80 years).

We then used a reference patient where all covariate values were prespecified and only patient race/ethnicity varied. Unlike the average patient approach above, this approach allowed for comparison of primary and secondary outputs across race/ethnicity categories, because all other covariates were held constant. The reference patient was chosen to be an unwell patient with a high likelihood of invasive ventilation: high-flow nasal cannula, heart rate 100 beats per minute, respiratory rate 25 breaths per minute, average age (66.5 years), female sex, no comorbidities, and admitted to the MIMIC-IV ICU. We varied inspired oxygen fraction from 0.50 to 1.0 to examine the influence of inspired oxygen fraction on predicted survival changes mediated by differences in invasive ventilation rates.

The model was implemented in R v4.0.3 and Stan and parallelized on the Niagara computing cluster.^39–42^ We used 4 parallel chains, with 250 iterations of warm-up and 250 iterations of sampling. Posterior distributions were summarized using mean and 95% credible interval (CrI).(Supplement)

### Sensitivity analyses

We repeated the analysis within MIMIC-IV and eICU cohorts. In the MIMIC-IV cohort analysis, we included insurance status and abnormal work-of-breathing as additional covariates. In the eICU cohort analysis, we included each hospital’s proportion of patients who were of non-white race/ethnicity. This covariate was included to test if differences by patient race/ethnicity could be explained by differences between hospitals that cared for many or few patients of non-white race/ethnicity.^13,43^

## Results

We studied 38,263 patients who met eligibility criteria, 48% (18,230) from eICU and 52% (20,033) from MIMIC-IV (Table 1, Figure S0). 46% were female with median age 68 years (interquartile range (IQR) 56-79). Asian patients comprised 2% (892), Black patients 11% (4,289), Hispanic patients 5% (1,964), and white patients 81% (31,118). Most patients were admitted between 2014 and 2016 (58%) to a medical-surgical ICU (66%) in a teaching hospital (63%) in the Northeast (57%). Patients were from 108 hospitals, 107 of which were from the eICU cohort.

**Table 1:** Cohort characteristics.

|  | Asian | Black | Hispanic | White |
| --- | --- | --- | --- | --- |
| **Total (N = 38,263)** | 892 | 4,289 | 1,964 | 31,118 |
| **Median age (interquartile range)** | 66 (55-79) | 61 (49-73) | 67 (52-80) | 69 (57-80) |
| **Sex (%)** |  |  |  |  |
| Female | 398 (45) | 2,193 (51) | 906 (46) | 14,218 (46) |
| **Comorbidities (%)** |  |  |  |  |
| CHF | 176 (20) | 1,135 (26) | 394 (20) | 7,508 (24) |
| COPD | 59 (7) | 559 (13) | 229 (12) | 5,396 (17) |
| Cancer | 223 (25) | 726 (17) | 287 (15) | 6,161 (20) |
| Dementia | 65 (7) | 440 (10) | 221 (11) | 3,293 (11) |
| **Year of ICU admission (%)** |  |  |  |  |
| 2008-2010 | 215 (24) | 1,127 (26) | 343 (17) | 6,651 (21) |
| 2011-2013 | 159 (18) | 535 (12) | 205 (10) | 4,039 (13) |
| 2014-2016 | 417 (47) | 2,375 (55) | 1,327 (68) | 17,913 (58) |
| 2017-2019 | 101 (11) | 252 (6) | 89 (5) | 2510 (8) |
| **Database (%)** |  |  |  |  |
| MIMIC-IV | 626 (70) | 2,887 (53) | 771 (39) | 16,356 (53) |
| eICU | 266 (30) | 2,005 (47) | 1,193 (61) | 14,762 (47) |
| **Region of hospital (%)** |  |  |  |  |
| Northeast | 649 (73) | 2,327 (54) | 776 (40) | 17,999 (58) |
| Midwest | 161 (18) | 1,203 (28) | 28 (1) | 9,631 (31) |
| West | 12 (1) | 7 (0) | 15 (1) | 280 (1) |
| South | 26 (3) | 602 (14) | 953 (49) | 2,548 (8) |
| Unknown | 44 (5) | 150 (3) | 192 (10) | 655 (2) |
| **Academic teaching hospital** | 669 (75) | 2,666 (62) | 938 (48) | 19,937 (64) |
| **Type of ICU** |  |  |  |  |
| Medical-surgical | 639 (72) | 3,744 (77) | 1,326 (68) | 20,211 (65) |
| Cardiac | 150 (17) | 774 (18) | 447 (23) | 7244 (23) |
| Neuro/trauma | 103 (12) | 374 (9) | 191 (10) | 3663 (12) |
| **Oxygen devices used** |  |  |  |  |
| Non-invasive ventilation | 49 (5) | 545 (13) | 422 (21) | 3764 (12) |
| High-flow nasal cannula | 29 (3) | 128 (3) | 46 (2) | 1780 (6) |
| Non-rebreather mask | 102 (11) | 389 (9) | 210 (11) | 3239 (10) |
| Face mask | 138 (15) | 436 (10) | 184 (9) | 4354 (14) |
| Nasal prongs | 835 (94) | 4061 (95) | 1840 (94) | 29457 (95) |
| **Time-weighted mean (95% interval)** |  |  |  |  |
| Heart rate (beats per minute) | 84 (55 to 119) | 85 (58 to 118) | 84 (59 to 120) | 82 (57 to 117) |
| Respiratory rate (breaths per minute) | 19.41 (13 to 29) | 20 (13 to 31) | 19 (13 to 30) | 19 (13 to 30) |
| Peripheral oxygen saturation | 98 (93 to 100) | 98 (93 to 100) | 98 (93 to 100) | 97 (92 to 100) |
| Inspired oxygen fraction | 0.28 (0.21 to 0.83) | 0.27 (0.21 to 0.79) | 0.33 (0.21 to 0.99) | 0.29 (0.21 to 0.81) |
| Glasgow Coma Scale | 15 (10 to 15) | 15 (11 to 15) | 15 (13 to 15) | 15 (12 to 15) |
| Vasopressor use (% of the total time) | 8 | 7 | 13 | 9 |

After incorporating time-varying covariates, re were 2,124,175 observations taken at a median interval of 60 minutes (95% interval 1 to 240). most common oxygen device was nasal prongs (used at least once by 95% of patients) while least common was high flow nasal cannula (used at least once by 5.2% of patients). Among clinical variables, time-weighted averages included: inspired oxygen fraction 0.29 (95% interval 0.21 to 0.82), peripheral oxygen saturation 97% (95% interval 92% to 100%), respiratory rate 19 breaths per minute (95% interval 13 to 30).

### Invasive ventilation rates

A total of 3,511 (9.2%) patients received invasive ventilation, of whom 92 were Asian, 333 Black, 106 Hispanic, and 2,980 white. Unadjusted 28-day invasive ventilation rate was 10.3% for Asian patients, 7.8% for Black patients, 5.4% for Hispanic patients, and 9.6% for white patients.

The cause-specific hazard ratio for invasive ventilation was lower in Asian (HR 0.82, CrI 0.70 to 0.95), Black (HR 0.78, CrI 0.71 to 0.86), and Hispanic (HR 0.70, CrI 0.61 to 0.79) patients as compared to white patients (Figure 1). The probability of a lower rate of invasive ventilation than white patients was greater than 0.99 for Asian, Black, and Hispanic patients. Hazard ratios for all covariates included are in the Supplement (Figure S4).

**Figure 1:**
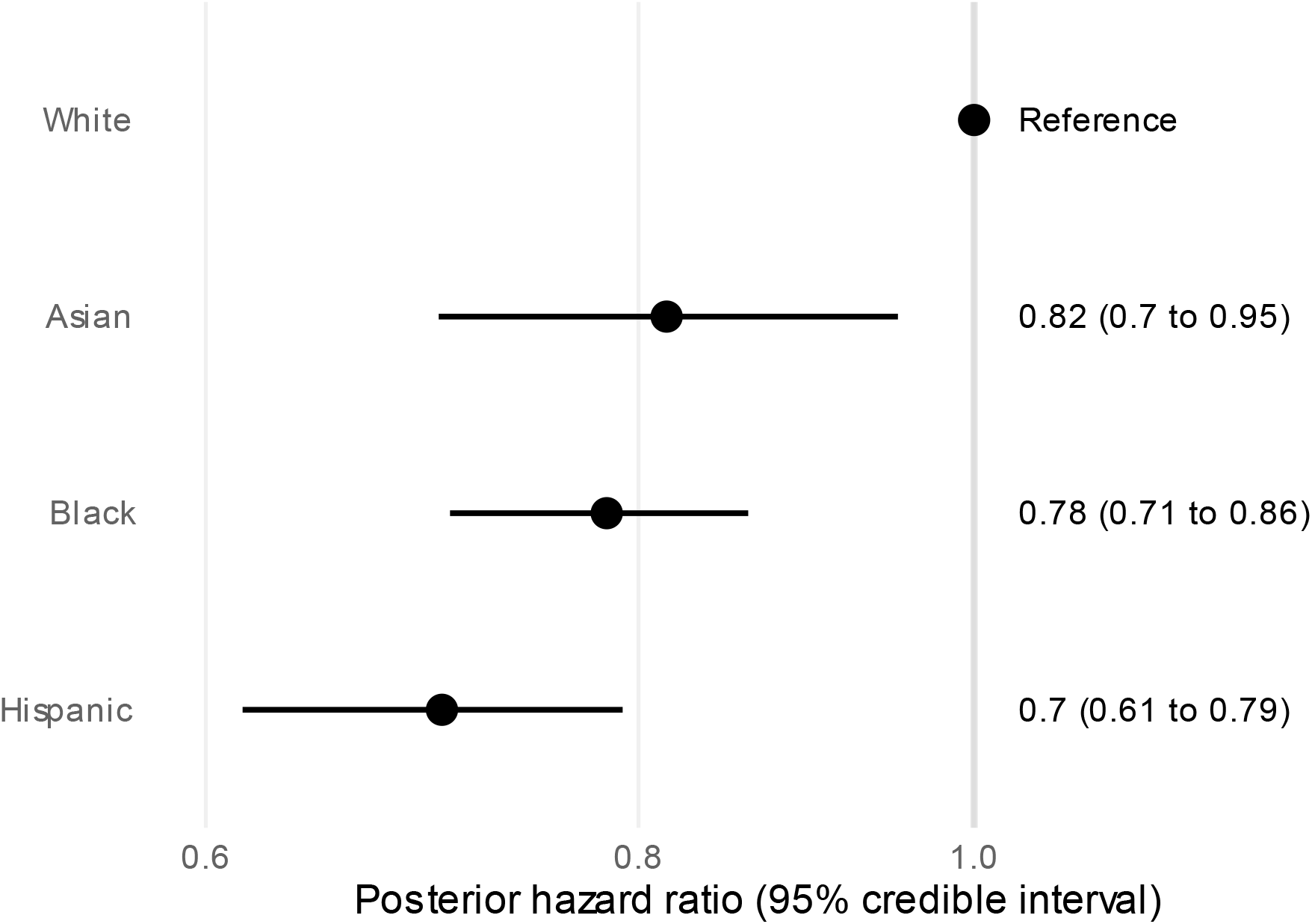
Cause-specific hazard ratios for invasive ventilation. This figure shows the posterior hazard ratios for the transition from oxygen therapy to invasive ventilation (x-axis) by patient race/ethnicity (y-axis). White race/ethnicity is the reference category. The x-axis scale is logarithmic. The posterior mean hazard ratios (with 95% credible intervals) are in the righthand column. The figure shows that Asian, Black, and Hispanic patients had a lower hazard of invasive ventilation than white patients, with the largest discrepancy seen in Hispanic patients.

### Hospital survival and impact of invasive ventilation rates

Death occurred in 2,869 (7.5%) patients, of whom 101 were Asian, 304 Black, 175 Hispanic, and 2,289 white. Unadjusted 28-day survival was 88.7% for Asian patients, 92.9% for Black patients, 91.1% for Hispanic patients, and 90.1% for white patients.

For the average Asian, Black, or Hispanic patient, the decreased invasive ventilation rate relative to white patients did not mediate a change in 28-day survival (Table 2). The probabilities that decreased rates of invasive ventilation mediated benefit for the average patient were 0.75 (Asian), 0.76 (Black), and 0.72 (Hispanic). Average patient covariates are available in Table S1. Findings were similar within sex and age subgroups (Tables S2, S3).

**Table 2:**
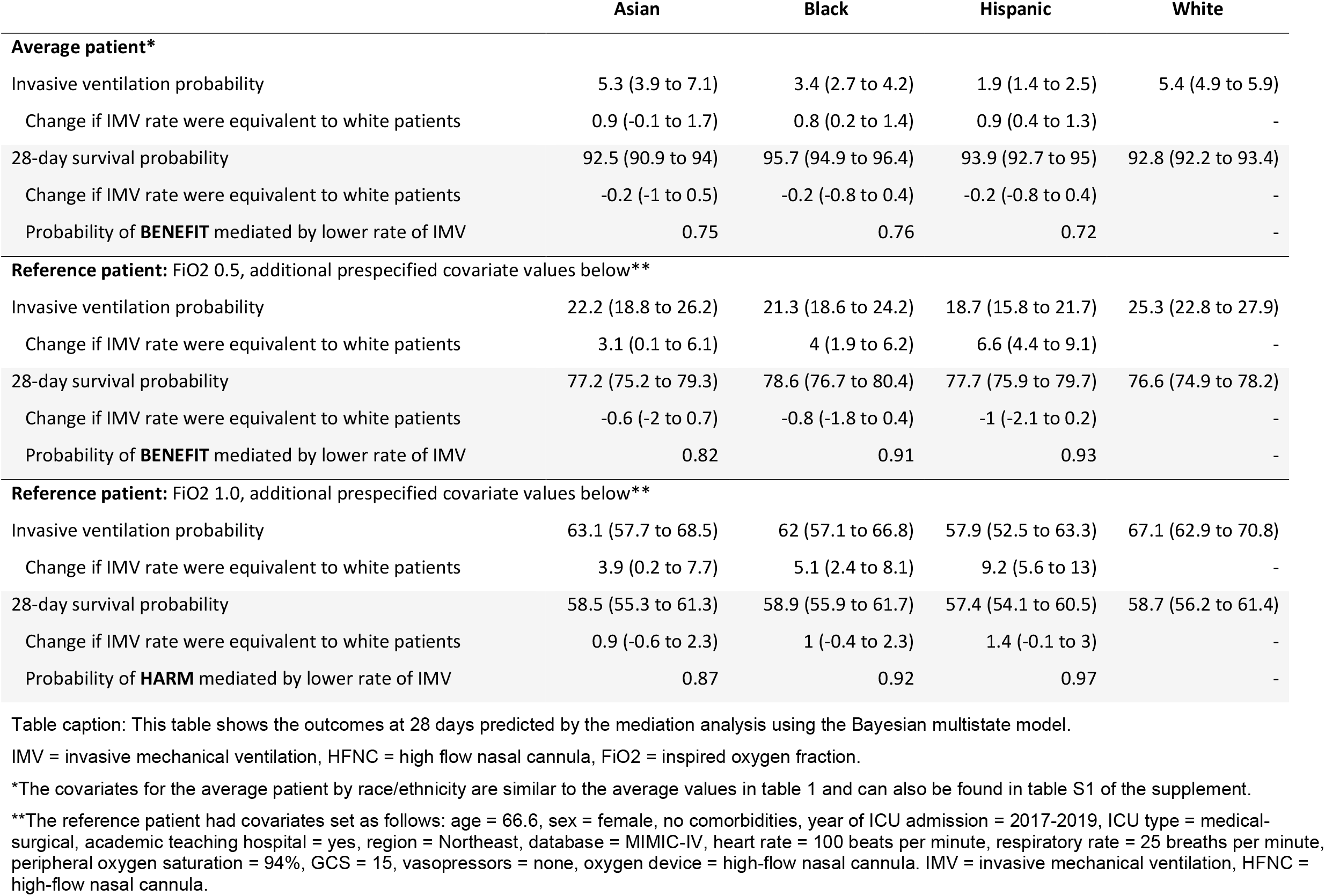
Mediation analysis outcomes at 28 days by patient race/ethnicity.

|  | Asian | Black | Hispanic | White |
| --- | --- | --- | --- | --- |
| **Average patient\*** |  |  |  |  |
| Invasive ventilation probability | 5.3 (3.9 to 7.1) | 3.4 (2.7 to 4.2) | 1.9 (1.4 to 2.5) | 5.4 (4.9 to 5.9) |
| Change if IMV rate were equivalent to white patients | 0.9 (-0.1 to 1.7) | 0.8 (0.2 to 1.4) | 0.9 (0.4 to 1.3) | - |
| 28-day survival probability | 92.5 (90.9 to 94) | 95.7 (94.9 to 96.4) | 93.9 (92.7 to 95) | 92.8 (92.2 to 93.4) |
| Change if IMV rate were equivalent to white patients | -0.2 (-1 to 0.5) | -0.2 (-0.8 to 0.4) | -0.2 (-0.8 to 0.4) | - |
| Probability of **BENEFIT** mediated by lower rate of IMV | 0.75 | 0.76 | 0.72 | - |
| **Reference patient: FiO2 0.5, additional prespecified covariate values below\*\*** |  |  |  |  |
| Invasive ventilation probability | 22.2 (18.8 to 26.2) | 21.3 (18.6 to 24.2) | 18.7 (15.8 to 21.7) | 25.3 (22.8 to 27.9) |
| Change if IMV rate were equivalent to white patients | 3.1 (0.1 to 6.1) | 4 (1.9 to 6.2) | 6.6 (4.4 to 9.1) | - |
| 28-day survival probability | 77.2 (75.2 to 79.3) | 78.6 (76.7 to 80.4) | 77.7 (75.9 to 79.7) | 76.6 (74.9 to 78.2) |
| Change if IMV rate were equivalent to white patients | -0.6 (-2 to 0.7) | -0.8 (-1.8 to 0.4) | -1 (-2.1 to 0.2) | - |
| Probability of **BENEFIT** mediated by lower rate of IMV | 0.82 | 0.91 | 0.93 | - |
| **Reference patient: FiO2 1.0, additional prespecified covariate values below\*\*** |  |  |  |  |
| Invasive ventilation probability | 63.1 (57.7 to 68.5) | 62 (57.1 to 66.8) | 57.9 (52.5 to 63.3) | 67.1 (62.9 to 70.8) |
| Change if IMV rate were equivalent to white patients | 3.9 (0.2 to 7.7) | 5.1 (2.4 to 8.1) | 9.2 (5.6 to 13) | - |
| 28-day survival probability | 58.5 (55.3 to 61.3) | 58.9 (55.9 to 61.7) | 57.4 (54.1 to 60.5) | 58.7 (56.2 to 61.4) |
| Change if IMV rate were equivalent to white patients | 0.9 (-0.6 to 2.3) | 1 (-0.4 to 2.3) | 1.4 (-0.1 to 3) | - |
| Probability of **HARM** mediated by lower rate of IMV | 0.87 | 0.92 | 0.97 | - |

Table caption: This table shows the outcomes at 28 days predicted by the mediation analysis using the Bayesian multistate model.
IMV = invasive mechanical ventilation, HFNC = high flow nasal cannula, FiO2 = inspired oxygen fraction.
\*The covariates for the average patient by race/ethnicity are similar to the average values in table 1 and can also be found in table S1 of the supplement.
\*\*The reference patient had covariates set as follows: age = 66.6, sex = female, no comorbidities, year of ICU admission = 2017-2019, ICU type = medical-surgical, academic teaching hospital = yes, region = Northeast, database = MIMIC-IV, heart rate = 100 beats per minute, respiratory rate = 25 breaths per minute, peripheral oxygen saturation = 94%, GCS = 15, vasopressors = none, oxygen device = high-flow nasal cannula. IMV = invasive mechanical ventilation, HFNC = high-flow nasal cannula.

For a prespecified reference patient, survival changes mediated by lower rates of invasive ventilation varied from probable modest benefit to probable modest harm (Figure 2). With inspired oxygen fraction of 0.5, lower rates of invasive ventilation mediated an increase in 28-day survival with probability 0.82 for Asian patients, 0.91 for Black patients, and 0.93 for Hispanic patients. However, the mean increase in survival was modest: 0.6% for Asian patients, 0.8% for Black patients, and 1.0% for Hispanic patients (Table 2). By contrast, with inspired oxygen fraction of 1.0, lower rates of invasive ventilation mediated a *decrease* in 28-day survival with probability 0.87 for Asian patients, 0.92 for Black patients, and 0.97 for Hispanic patients. The corresponding mean survival decrease was again modest: 0.9% for Asian patients, 1.0% for Black patients, and 1.4% for Hispanic patients (Table 2).

**Figure 2:**
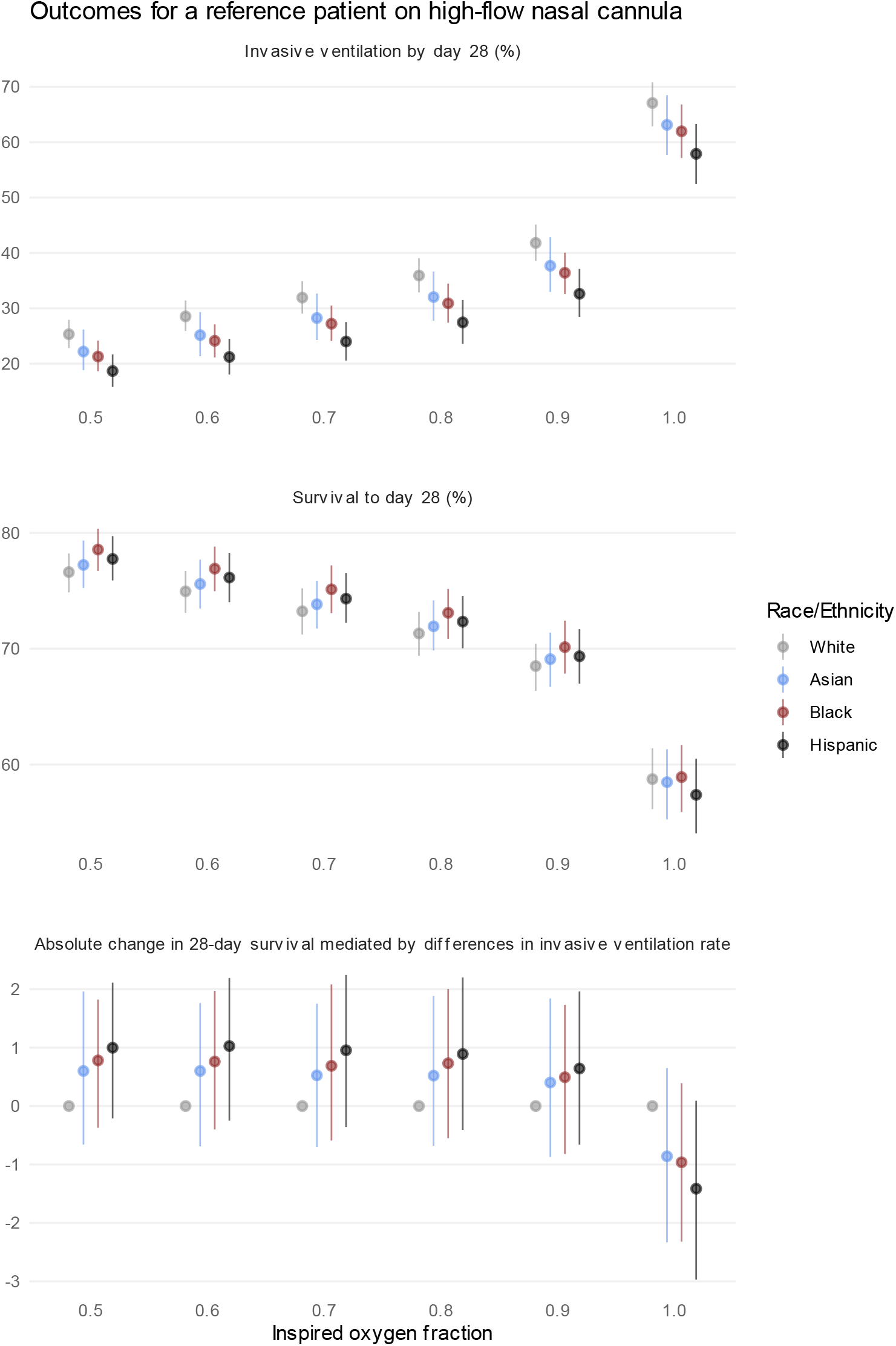
Predicted outcomes for a patient on high-flow nasal cannula. This figure shows the mediation analysis outcomes (y-axis) by patient race/ethnicity (colors) for the reference patient at different inspired oxygen fractions (x-axis) ranging from 0.5 to 1.0. The top panel shows the probability of invasive ventilation by 28 days, the middle panel shows the probability of 28-day survival, and the bottom panel shows the absolute change in 28-day survival mediated by differences in the rate of invasive ventilation. Other than inspired oxygen fraction, the reference patient had covariates set as follows: age = 67, sex = female, no comorbidities, year of ICU admission = 2017-2019, ICU type = medical-surgical, academic teaching hospital = yes, region = Northeast, database = MIMIC-IV, heart rate = 100 beats per minute, respiratory rate = 25 breaths per minute, peripheral oxygen saturation = 94%, GCS = 15, vasopressors = none, oxygen device = high-flow nasal cannula.

### Sensitivity analyses and model diagnostics

Results were consistent in a sensitivity analysis restricted to the MIMIC-IV cohort that included additional covariates of insurance status and time-varying work of breathing. In a sensitivity analysis restricted to the eICU cohort, Asian, Black, and Hispanic race/ethnicity remained associated with a lower rate of invasive ventilation (Figure S11). After adding a hospital-level covariate corresponding to the proportion of non-white patients being cared for in that hospital’s ICU, the hazard ratio of invasive ventilation associated with Asian and Black race/ethnicity was slightly increased, and the hazard ratio associated with Hispanic race/ethnicity was markedly increased: Asian 0.92 (CrI 0.76 to 1.00), Black 0.82 (CrI 0.76 to 1.0), and Hispanic 1.00 (CrI 0.82 to 1.20) (Figure S12). The hazard ratio for invasive ventilation associated with being cared for in a hospital that had a 10% higher (absolute percent change) proportion of non-white ICU patients was 0.75 (CrI 0.71 to 0.79).

The Bayesian multistate model used to calculate cause-specific hazard ratios and perform mediation analysis showed good convergence and model diagnostics. Model diagnostics and hazard ratios for all transitions are in the supplement.

## Discussion

This cohort study of 38,263 patients receiving supplemental oxygen in ICU found lower rates of invasive ventilation in Asian, Black, or Hispanic patients as compared to white patients, despite adjustment for baseline covariates and time-varying severity. For the average patient, lower rates were not associated with harm. For a prespecified reference patient receiving oxygen via high-flow nasal cannula, the change in 28-day survival mediated by lower rates of invasive ventilation ranged from probable modest benefit at inspired oxygen fractions of 0.9 or less to probable modest harm at an inspired oxygen fraction of 1.0.

These findings differ from prior research that shows increased rates of invasive ventilation in non-white patients compared with white patients. Studies of Medicare beneficiaries with advanced dementia and COPD patients in the Veteran’s Affairs health system found an increased invasive ventilation rate in Black as opposed to white patients.^18,19^ Research in Canada and the USA showed an increased rate of invasive ventilation at end-of-life for Asian, Black, and Hispanic decedents as compared to white decedents.^20–23^ Results are conflicting in patients with COVID-19 pneumonia.^24–27^ No studies incorporated adjustment for time-varying severity, meaning that prior observed differences in rates of invasive ventilation by race/ethnicity could be explained by greater time-varying clinical severity.

Pulse oximeters are known to overestimate arterial saturation in patients with darker skin pigment, but this is unlikely to explain our results.^7,9,44,45^ In our analysis we incorporated peripheral but not arterial oxygen saturation, but hidden hypoxemia has been associated with greater severity of illness and hospital mortality.^46^ This suggests that adjustment for peripheral but not arterial saturation would bias results towards a higher rate of invasive ventilation, the opposite direction from our findings.

Lack of information on socioeconomic position could explain our results. Lower socioeconomic status is associated with Black or Hispanic race/ethnicity in the USA.^47^ Lower invasive ventilation rates by race/ethnicity could be explained by patient, family, or clinician reluctance to use more expensive treatments in patients with lower socioeconomic position. However, our sensitivity analysis in the MIMIC-IV cohort including an insurance status covariate showed similar results, and other research has shown that residing in a lower socioeconomic neighbourhood is associated with similar or higher rates of invasive ventilation among patients with COVID-19.^26,27^

Our results suggest that invasive ventilation rate differences could be explained by variation in hospital practice associated with the proportion of patients of non-white race/ethnicity cared for in each hospital’s ICUs. In the USA, patients of different race/ethnicities are clustered at hospital level, with some hospitals caring for a disproportionately high or low proportion of Black or Hispanic patients.^13^ In one study of ICU admission rates, hospital-level variation offered a numerical explanation for differences in ICU admission rates by patient race/ethnicity at population level.^43^

Another study of end-of-life care also found that within the same hospital, Black patients received less invasive ventilation than white patients, but across all hospitals, Black patients received more invasive ventilation than white patients.^48^ This supports our findings that differences in invasive ventilation can be explained by both within-hospital and across-hospital variability, highlighting the multilevel impact of systemic racism.

Bias in clinical decision-making or recognition of deterioration is a potential explanation for the findings. Such bias could result from overt racism or implicit bias.^14,49^ Awareness of implicit bias could offer some benefit, especially if education efforts incorporate expert guidance.^50,51^ However, bias-reduction initiatives may not generate durable improvement.^52^ To improve equity of care for patients with respiratory failure, we will require additional strategies, such as physiologic thresholds for invasive ventilation.

Our results are consistent with the existence of optimal physiologic thresholds for invasive ventilation, because they imply that invasive ventilation has benefit for patients with sufficient severity of illness, and otherwise may cause harm. This interpretation is supported by prior work showing a mortality benefit with invasive ventilation for patients with acute respiratory distress syndrome and an arterial-to-inspired oxygen ratio less than 150 mmHg, or patients with COVID-19 pneumonia and higher baseline severity as measured by Sequential Organ Failure Assessment (SOFA) score.^53,54^ The optimal physiologic thresholds for invasive ventilation are unknown^55^, but our analysis suggests that finding disease-specific thresholds for invasive ventilation could improve outcomes for white patients at lower severity, and for Asian, Black, and Hispanic patients at higher severity.

Important limitations of this study include a low proportion of patients with Asian, Black, or Hispanic race/ethnicity, a lack of international data, validity of the race/ethnicity data, parametric model specification, and unmeasured confounding. Future research should aim to replicate these findings in patient populations with a higher proportion of Asian, Black, and Hispanic patients, and in additional populations outside the USA. Race/ethnicity data used in this study was gathered in the course of hospital admission, and self-identified race or ethnicity may not always match the database value. Further, the included categories of race/ethnicity encompass large, heterogeneous populations, and muddle together distinct concepts of race and ethnicity. Replication of these results using more robust data collection with nuanced self-categorization will be important. Regarding model specification, the multistate model uses several parametric components, which means mediation analysis results may be biased if models were mis-specified. Last, unmeasured confounding could explain these results. Additional studies incorporating more comprehensive social determinants of health data would help clarify the association between patient race/ethnicity and invasive ventilation use.

The strengths of this study are its sample size, methodology, and focus on an actionable change in management. A sample size of 38,263 patients allowed for a large set of known confounding variables and adjustment for time-varying severity. The methodology was novel both because it adjusted for time-varying severity and it incorporated a mediation analysis focusing on downstream clinical impact of any observed association.^38^ We focused on invasive ventilation rates because they present an actionable pathway to more equitable care, via use of physician self-reflection and physiologic thresholds for invasive ventilation to reduce the influence of patient race/ethnicity on clinical decision-making.

## Conclusions

Asian, Black, and Hispanic patients had lower rates of invasive ventilation than white patients, after controlling for time-varying severity and baseline confounders. Although change in survival mediated by lower rates of invasive ventilation ranged from slight benefit at lower inspired oxygen fractions to slight harm at inspired oxygen fraction of 1.0, absolute change was consistently less than 1.5%. Further research on optimal thresholds for invasive ventilation, increased funding support for hospitals with minoritized patient populations, and adoption of strategies to mitigate implicit bias may help reduce differences in rates of invasive ventilation by patient race/ethnicity.

## Data Availability

All data are available through the Physionet repository at physionet.org.

https://physionet.org/content/mimiciv/2.0/

https://physionet.org/content/eicu-crd/2.0/

## Acknowledgements

We thank the following for helpful comments: Martin Urner.

