## Supplement for "Association between patient race/ethnicity and invasive ventilation in hypoxemic respiratory failure"

Fred M. Abdelmalek,

Federico Angriman,

Julie Moore,

Kuan Liu,

Lisa Burry,

Laleh Seyyed-Kalantari,

Sangeeta Mehta,

Judy Gichoya,

Leo Anthony Celi,

George Tomlinson,

Michael Fralick,

Christopher J. Yarnell

October 27, 2022

#### STROBE checklist

| | Item No | Recommendation | Page No |
|---|---|---|---|
| **Title and abstract** | 1 | (*a*) Indicate the study's design with a commonly used term in the title or the abstract | 5 |
| | | (*b*) Provide in the abstract an informative and balanced summary of what was done and what was found | 5 |
| **Introduction** | | | |
| Background/rationale | 2 | Explain the scientific background and rationale for the investigation being reported | 6-7 |
| Objectives | 3 | State specific objectives, including any prespecified hypotheses | 7 |
| **Methods** | | | |
| Study design | 4 | Present key elements of study design early in the paper | 7-8 |
| Setting | 5 | Describe the setting, locations, and relevant dates, including periods of recruitment, exposure, follow-up, and data collection | 7-8 |
| Participants | 6 | (*a*) Give the eligibility criteria, and the sources and methods of selection of participants. Describe methods of follow-up | 7-8 |
| | | (*b*) For matched studies, give matching criteria and number of exposed and unexposed | |
| Variables | 7 | Clearly define all outcomes, exposures, predictors, potential confounders, and effect modifiers. Give diagnostic criteria, if applicable | 8 |
| Data sources/ measurement | 8* | For each variable of interest, give sources of data and details of methods of assessment (measurement). Describe comparability of assessment methods if there is more than one group | 8, supplement |
| Bias | 9 | Describe any efforts to address potential sources of bias | 9-11 |
| Study size | 10 | Explain how the study size was arrived at | 7 |
| Quantitative variables | 11 | Explain how quantitative variables were handled in the analyses. If applicable, describe which groupings were chosen and why | 8 |
| Statistical methods | 12 | (*a*) Describe all statistical methods, including those used to control for confounding | 9 |
| | | (*b*) Describe any methods used to examine subgroups and interactions | 9 |
| | | (*c*) Explain how missing data were addressed | Supp 13 |
| | | (*d*) If applicable, explain how loss to follow-up was addressed | 9 |
| | | (*e*) Describe any sensitivity analyses | 11 |
| **Results** | | | |
| Participants | 13* | (a) Report numbers of individuals at each stage of study—eg numbers potentially eligible, examined for eligibility, confirmed eligible, included in the study, completing follow-up, and analysed | 11 |
| | | (b) Give reasons for non-participation at each stage | Supp 17 |

| | | | |
|---|---|---|---|
| | | (c) Consider use of a flow diagram | Supp 17 |
| Descriptive data | 14* | (a) Give characteristics of study participants (eg demographic, clinical, social) and information on exposures and potential confounders | 11 |
| | | (b) Indicate number of participants with missing data for each variable of interest | Supp 17 |
| | | (c) Summarise follow-up time (eg, average and total amount) | N/A |
| Outcome data | 15* | Report numbers of outcome events or summary measures over time | 11-12 |
| Main results | 16 | (*a*) Give unadjusted estimates and, if applicable, confounder-adjusted estimates and their precision (eg, 95% confidence interval). Make clear which confounders were adjusted for and why they were included | 12-13 |
| | | (*b*) Report category boundaries when continuous variables were categorized | N/A |
| | | (*c*) If relevant, consider translating estimates of relative risk into absolute risk for a meaningful time period | 13 |
| Other analyses | 17 | Report other analyses done—eg analyses of subgroups and interactions, and sensitivity analyses | 13 |
| **Discussion** | | | |
| Key results | 18 | Summarise key results with reference to study objectives | 14 |
| Limitations | 19 | Discuss limitations of the study, taking into account sources of potential bias or imprecision. Discuss both direction and magnitude of any potential bias | 16-17 |
| Interpretation | 20 | Give a cautious overall interpretation of results considering objectives, limitations, multiplicity of analyses, results from similar studies, and other relevant evidence | 18 |
| Generalisability | 21 | Discuss the generalisability (external validity) of the study results | 16-17 |
| **Other information** | | | |
| Funding | 22 | Give the source of funding and the role of the funders for the present study and, if applicable, for the original study on which the present article is based | 2 |

### Cohort creation

Cohort construction was carried out in a similar fashion to a recent study of whether patients receive invasive ventilation after they meet physiologic thresholds. All variable transformation choices are the same as in that cohort.(1)

#### Inclusion criteria

Patients were included if they met all of the following criteria:

1) Adults

2) Received noninvasive supplemental oxygen within the first 24h of ICU admission

Patients were excluded if they met any of the following criteria:

1) Invasively ventilated prior to the first observation of noninvasive supplemental oxygen
2) Race/ethnicity recorded as something other than Black, Asian, Hispanic, or White.
3) Admission classified as "SURGICAL SAME DAY ADMISSION" or "ELECTIVE" in the MIMIC-IV database (intended to filter out patients admitted from the operating room)
4) Tracheostomy charted within the first 7 days of ICU admission
5) Goals of care charted as not for invasive ventilation in the MIMIC-IV cohort

In MIMIC-IV, the additional options for race/ethnicity were American Indian/Alaska Native, unknown, unable to obtain, and other. In eICU, the additional options were missing, other/unknown, and Native American. Patients with American Indian/Alaska Native or Native American race/ethnicity were not included due to small numbers.

#### Specific variables

Below we document the decision-making for extracting and transforming specific variables.

##### Care unit

The different types of care units were collapsed into three categories: Medical-Surgical, Cardiac, and Neuro-Trauma. From MIMIC-IV, "Coronary Care Unit (CCU)” and "Cardiac Vascular Intensive Care Unit (CVICU)" were cardiac; "Medical Intensive Care Unit (MICU)”, "Surgical Intensive Care Unit (SICU)", "Medical/Surgical Intensive Care Unit (MICU/SICU)" were Medical-Surgical; "Neuro Surgical Intensive Care Unit (Neuro SICU)", "Neuro Stepdown", "Neuro Intermediate" and "Trauma SICU (TSICU)" were Neuro-Trauma. were collapsed into three categories: Medical-Surgical, Cardiac, and Neuro-Trauma. From eICU, "CTICU","Cardiac ICU",

"CCU-CTICU","CSICU" were Cardiac; "Neuro ICU" was Neuro-Trauma, and the remainder were Medical-Surgical.

#### Fraction of inspired oxygen

Fraction of inspired oxygen was taken directly from the corresponding field where available. If no charted fraction of inspired oxygen was available, and the patient was receiving oxygen by non-rebreather mask, face mask, or nasal prongs, then the oxygen flow was used to estimate the fraction of inspired oxygen by a validated equation(16): 21% + oxygen flow rate in L/min*3. For analysis, fraction of inspired oxygen was transformed from the interval [21,100] to the real line (-infinity, infinity) with the logit function, centered at the mean, and scaled by the standard deviation.

#### Work of breathing

In the MIMIC-IV data there was information about work of breathing. This variable was composed from several different fields relating to the pattern of respiration (chart event IDs 229322, 223990, 229323). Patterns described as 'Dyspneic', 'Labored', 'Shallow', 'Apneic', 'Agonal', 'Discoordinate', 'Gasping efforts', 'Prolonged exhalation', 'Shallow', 'Irregular', 'Nasal flaring', 'Cheyne-Stokes', 'Accessory muscle use/retractions', 'Frequent desaturation episodes', 'Inability to speak in full sentences', and 'Active exhalation' were classified as abnormal. Normal observations included 'Regular' or 'Normal'. The chart event ID 229323 was labeled the “Current Dyspnea Assessment” and recorded dyspnea on a scale from 0 to 10. Dyspnea levels of 'Moderate - 4', 'Moderate - 5', 'Moderate - 6', 'Moderate - 7', 'Severe - 8', 'Severe - 9', or

'Severe - 10' were classified as abnormal, while dyspnea levels of 'None - 0', 'Mild - 1', 'Mild - 2', or 'Mild - 3' were classified as normal.

#### Proportion of patients of non-white race/ethnicity (eICU sensitivity analysis)

In the eICU sensitivity analysis we incorporated the proportion of patients cared for at each hospital that were of non-White race/ethnicity (race/ethnicities recorded as Black, Asian, Hispanic, or Native American). This quantity ranged from 0 to 1. We transformed it by centering at the median (0.12) and multiplying by 10, such that an increase of 0.1 in the proportion of patients of non-white race/ethnicity at a hospital corresponded to one odds ratio of increase in the hazard of invasive ventilation.

#### Comorbidities

We incorporated the presence or absence of four comorbidities: cancer, congestive heart failure (CHF), chronic obstructive pulmonary disease (COPD), and dementia. In MIMIC-IV we used ICD codes to identify dementia (F0 to F3 or 290), cancer (C0 to C9, D0 to D4, 14x to 19x, 20x to 23x), COPD (20500), and CHF (496, 49121, J449, J441, J440, J439). In eICU used the structured text field of *pasthistoryvalue* and classified a patient as having CHF if “CHF” appeared, COPD if “COPD” appeared, dementia if “dementia” appeared, and cancer if “Cancer” appeared.

#### Other variables

To facilitate convergence of the model, continuous variables were transformed. Age was centered at the mean and scaled by the standard deviation. For heart rate and respiratory rate, we used the logarithm of the observed value, centered at the mean (of the logarithm of the data), and scaled by the standard deviation. For peripheral saturation and Glasgow Coma Scale we transformed the interval data (intervals [0,100] and [3,15] respectively) to the real line using logit functions, then centered at the mean and scaled by standard deviation.

#### Bayesian multistate model

We used a continuous time Bayesian multistate model. Possible states included oxygen therapy, invasive ventilation, ICU discharge, and death (Figure S1). We did not allow for two-way transitions, which means that the invasive ventilation state would be more accurately labelled as “invasive ventilation at least once”, and the ICU discharge state would be more accurately labelled as “ICU discharge at least once.” However, we could not accommodate the complexity that would be introduced by two-way transitions, and we felt that the clinical scenario of reintubation was not the same as the scenario of *de novo* invasive ventilation.

##### Model structure

For all transitions, we used the following set of baseline confounders: patient race/ethnicity (White, Black, Asian, Hispanic), age, sex, comorbidities (COPD, CHF, cancer, dementia), admission year group, ICU type, teaching hospital status, hospital region, and database.

For the transitions from oxygen therapy to other states, we incorporated time-varying covariates and used an exponential function for the baseline hazard. The time-varying covariates were heart rate, respiratory rate, peripheral oxygen saturation, inspired oxygen fraction, vasopressor use, and oxygen device. These were intended to account for any differences in physiologic severity or evolution across patients of different race/ethnicity. This was particularly important given that our cohort consisted of patients who had been admitted to the ICU. We did not want the impact of bias at the level of ICU admission (for example, if patients of a given race/ethnicity were admitted to ICU “later” or at higher severity) to confound the results. The number of time-varying covariates was high, generating a table of more than 2 million rows for the approximately 40,000 patients (~50 observations per patient). We chose an exponential function for the baseline hazard to avoid model identification issues between the time-varying covariates and a changing hazard over time.

For all other transitions, we used an accelerated failure time model with Weibull hazard. These transitions all originated from the invasive ventilation or ICU discharge states and so we also included a covariate of the logarithm of the time of entry into that state.

#### Mediation analysis

To generate estimates of the change in survival mediated by differences in time-to-invasive ventilation, we performed Monte Carlo integration using the *generated quantities* function of Stan. We used the posterior distribution of the model parameters to simulate each patient’s trajectory from initial eligibility to 28-day outcome, keeping track of whether or not invasive ventilation occurred, and whether or not the patient survived. For each sample of the posterior distribution of model parameters, we averaged the probability of invasive ventilation and survival over 10,000 replicates of the patient in question (average patient or reference patient), in order to minimize Monte Carlo error.

The specific mediation quantity (change in absolute survival mediated by the difference in time-to-invasive ventilation) was calculated in the counterfactual manner, inspired by Valeri et al and in accordance with recommendations by Lapointe-Shaw et al.(2,3) In essence, for each non-white race/ethnicity, we compared the survival probability with the usual time-to-invasive ventilation to the survival probability with the time-to-invasive ventilation distribution of white patients. This amounted to simulating trajectories with the usual hazard ratios for each transition, and simulating trajectories with the time-to-invasive ventilation hazard ratio corresponding to race/ethnicity changed from the particular non-white race/ethnicity value to the reference value for white patients – and all other hazard ratios, for both the transition to invasive ventilation and all other transitions, remained the same.

For the average and reference patients we held the time-varying covariate values constant. An alternative approach would have been to build a model for the time-varying covariates, but this is a complex modeling task that would introduce additional limitations. We left this more complicated approach as an avenue for future research.

#### Computational details

The models were programmed in Stan using 250 warmup iterations, 250 sampling iterations, and 4 parallel chains. We used within-in chain parallelization and performed the computation using the Niagara computer cluster from the Digital Research Alliance Canada. All code is available at https://doi.org/10.5281/zenodo.7267580.

#### Model diagnostics

The model converged with satisfactory estimated Bayesian fraction of missing information, no divergences in any chain, and no samples hitting the maximum tree depth.(4,5) Example trace plots are shown in Figure S1. We further inspected the correlation plot between posterior parameter values, because of the large number of included predictors. This did not support any meaningful collinearity between the primary study parameters (race/ethnicity) and other variables.

### Tables

#### Table 0: Missing data

| Variable | Missing Proportion | Missing Count |
|---|---|---|
| Age | 0 | 0 |
| Sex | 0 | 0 |
| CHF | 0 | 0 |
| COPD | 0 | 0 |
| Cancer | 0 | 0 |
| Dementia | 0 | 0 |
| Admission year | 0 | 0 |
| ICU type | 0 | 0 |
| Region | 0 | 0 |
| Database | 0 | 0 |
| Heart rate | 0.013 | 29512 |
| Respiratory rate | 0.012 | 27363 |
| Peripheral saturation | 0.013 | 30554 |
| Inspired oxygen fraction | 0.026 | 60341 |
| Glasgow Coma Scale | 0.107 | 248422 |
| Vasopressor use | 0 | 0 |
| Oxygen device | 0 | 0 |

This table shows the proportion and count of missing data after finishing cohort construction (see Figure S0) and performing forward-fill imputation. The remaining missing data shown here was imputed with the mean. Because all covariates were centered at 0 for the modeling, imputing the mean was equivalent to making that row neutral with respect to whether that coefficient increased or decreased the probability of that transition.

#### Table S1: Characteristics of the average patients for mediation analysis

| | White | Black | Hispanic | Asian |
|---|---|---|---|---|
| **Age (years)** | 67.27 | 60.43 | 64.64 | 64.74 |
| **Sex** | | | | |
| Female | 0.46 | 0.51 | 0.46 | 0.45 |
| **Comorbidities** | | | | |
| CHF | 0.24 | 0.26 | 0.20 | 0.20 |
| COPD | 0.17 | 0.13 | 0.12 | 0.07 |
| Cancer | 0.20 | 0.17 | 0.15 | 0.25 |
| Dementia | 0.11 | 0.10 | 0.11 | 0.07 |
| **Year of ICU admission** | | | | |
| 2008-2010 | 0.21 | 0.26 | 0.17 | 0.24 |
| 2011-2013 | 0.13 | 0.12 | 0.10 | 0.18 |
| 2014-2016 | 0.58 | 0.55 | 0.68 | 0.47 |
| 2017-2019 | 0.08 | 0.06 | 0.05 | 0.11 |
| **Database** | | | | |
| MIMIC-IV | 0.53 | 0.53 | 0.39 | 0.70 |
| eICU | 0.47 | 0.47 | 0.61 | 0.30 |
| **Region of hospital** | | | | |
| Northeast | 0.58 | 0.54 | 0.40 | 0.73 |
| Midwest | 0.31 | 0.28 | 0.01 | 0.18 |
| West | 0.01 | 0.00 | 0.01 | 0.01 |
| South | 0.08 | 0.14 | 0.49 | 0.03 |
| Unknown | 0.02 | 0.03 | 0.10 | 0.05 |
| **Academic teaching hospital** | 0.64 | 0.62 | 0.48 | 0.75 |
| **Type of ICU** | | | | |
| Medical-surgical | 0.65 | 0.73 | 0.68 | 0.72 |
| Cardiac | 0.23 | 0.18 | 0.23 | 0.17 |
| Neuro/trauma | 0.12 | 0.09 | 0.10 | 0.12 |
| **Oxygen devices** | | | | |
| Room air | 0.05 | 0.04 | 0.04 | 0.03 |
| Nasal prongs | 0.80 | 0.82 | 0.79 | 0.83 |
| Facemask | 0.05 | 0.04 | 0.03 | 0.07 |
| Non-rebreather mask | 0.03 | 0.03 | 0.03 | 0.04 |
| Non-invasive ventilation | 0.05 | 0.06 | 0.09 | 0.02 |
| High-flow nasal cannula | 0.02 | 0.01 | 0.01 | 0.02 |
| **Time-varying covariates** | | | | |
| Heart rate (beats per minute) | 82.01 | 84.39 | 83.17 | 83.91 |
| Respiratory rate (breaths per minute) | 19.20 | 19.55 | 19.19 | 19.30 |
| Peripheral oxygen saturation | 96.64 | 97.76 | 97.51 | 97.42 |
| Inspired oxygen fraction | 30.72 | 28.68 | 35.20 | 30.06 |
| Glasgow Coma Scale | 14.87 | 14.88 | 14.94 | 14.73 |
| Vasopressor use (proportion of time) | 0.09 | 0.07 | 0.13 | 0.08 |

#### Table S2: Outcomes at 28 days by subgroup (sex)

| | Asian | Black | Hispanic | White |
|---|---|---|---|---|
| **Average female patient** | | | | |
| Invasive ventilation probability | 4.9 (3.6 to 6.7) | 3.5 (2.7 to 4.3) | 1.7 (1.2 to 2.2) | 5.2 (4.7 to 5.8) |
| Change with time-to-IMV equivalent to white patients | 0.8 (-0.2 to 1.7) | 0.8 (0.2 to 1.4) | 0.8 (0.3 to 1.1) | - |
| 28-day survival probability | 92.1 (90.5 to 93.6) | 95.1 (94.3 to 95.9) | 93.1 (91.6 to 94.5) | 91.9 (91.2 to 92.5) |
| Change with time-to-IMV equivalent to white patients | -0.3 (-0.9 to 0.4) | -0.2 (-0.8 to 0.3) | -0.2 (-0.8 to 0.5) | - |
| **Average male patient** | | | | |
| Invasive ventilation probability | 5.6 (4.1 to 7.5) | 3.3 (2.6 to 4.1) | 2 (1.5 to 2.7) | 5.5 (5 to 6) |
| Change with time-to-IMV equivalent to white patients | 0.9 (-0.1 to 1.8) | 0.8 (0.2 to 1.4) | 1 (0.4 to 1.4) | - |
| 28-day survival probability | 92.9 (91.4 to 94.2) | 96.2 (95.4 to 96.8) | 94.4 (93.3 to 95.4) | 93.5 (92.9 to 94) |
| Change with time-to-IMV equivalent to white patients | -0.3 (-1 to 0.4) | -0.2 (-0.8 to 0.3) | -0.2 (-0.8 to 0.4) | - |

#### Table S3: Outcomes at 28 days by subgroup (age)

| | Asian | Black | Hispanic | White |
|---|---|---|---|---|
| **Average patient with age < 50 years** | | | | |
| Invasive ventilation probability | 4.9 (3.5 to 6.5) | 3.5 (2.7 to 4.3) | 3 (2.2 to 4) | 5.7 (5.1 to 6.3) |
| Change with time-to-IMV equivalent to white patients | 0.8 (-0.1 to 1.7) | 0.8 (0.2 to 1.4) | 1.4 (0.8 to 2) | - |
| Survival probability | 96 (95 to 96.9) | 97.4 (96.9 to 97.9) | 96.3 (95.4 to 97.1) | 96 (95.5 to 96.5) |
| Change with time-to-IMV equivalent to white patients | -0.2 (-0.7 to 0.4) | -0.2 (-0.6 to 0.3) | -0.2 (-0.7 to 0.3) | - |
| **Average patient with age 50 to 65 years** | | | | |
| Invasive ventilation probability | 5.7 (4.2 to 7.6) | 3.4 (2.7 to 4.2) | 2.4 (1.8 to 3.2) | 5.6 (5.1 to 6.1) |
| Change with time-to-IMV equivalent to white patients | 0.9 (-0.2 to 1.8) | 0.8 (0.2 to 1.4) | 1.1 (0.6 to 1.6) | - |
| Survival probability | 93.3 (91.8 to 94.6) | 96 (95.3 to 96.6) | 94.5 (93.3 to 95.6) | 94 (93.4 to 94.6) |
| Change with time-to-IMV equivalent to white patients | -0.3 (-0.9 to 0.4) | -0.2 (-0.8 to 0.3) | -0.3 (-0.9 to 0.4) | - |
| **Average patient with age 65 to 80 years** | | | | |
| Invasive ventilation probability | 4.4 (3.2 to 5.9) | 3.2 (2.5 to 4) | 1.6 (1.2 to 2.2) | 5.1 (4.7 to 5.6) |
| Change with time-to-IMV equivalent to white patients | 0.7 (-0.1 to 1.5) | 0.8 (0.1 to 1.3) | 0.8 (0.3 to 1.2) | - |
| Survival probability | 91.9 (90.3 to 93.4) | 94.4 (93.5 to 95.3) | 92.8 (91.3 to 94.1) | 92.2 (91.6 to 92.7) |
| Change with time-to-IMV equivalent to white patients | -0.3 (-1 to 0.5) | -0.2 (-0.9 to 0.4) | -0.2 (-0.9 to 0.6) | - |
| **Average patient with age > 80 years** | | | | |
| Invasive ventilation probability | 6.5 (4.7 to 8.7) | 3.7 (2.9 to 4.6) | 1.1 (0.8 to 1.5) | 5.2 (4.7 to 5.7) |
| Change with time-to-IMV equivalent to white patients | 1.1 (-0.1 to 2.1) | 0.9 (0.3 to 1.5) | 0.5 (0.2 to 0.9) | - |
| Survival probability | 87.4 (85 to 89.6) | 91.8 (90.5 to 93) | 91 (89.1 to 92.6) | 89.3 (88.5 to 90.1) |
| Change with time-to-IMV equivalent to white patients | -0.3 (-1.3 to 0.6) | -0.3 (-1.1 to 0.5) | -0.1 (-0.9 to 0.7) | - |

### Figures

#### Figure S0: Cohort flow diagram

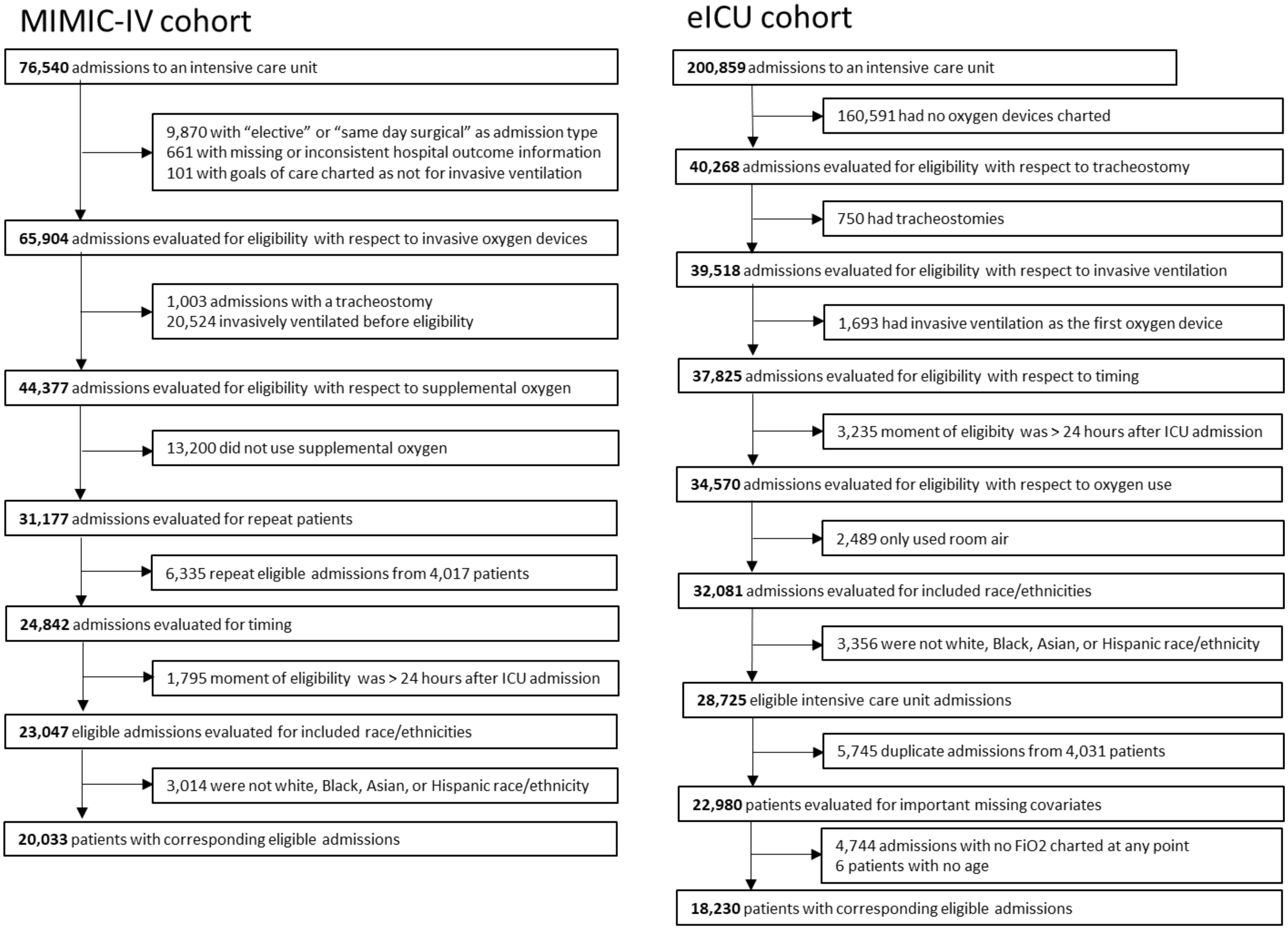

#### Figure S1: Bayesian multistate model diagram

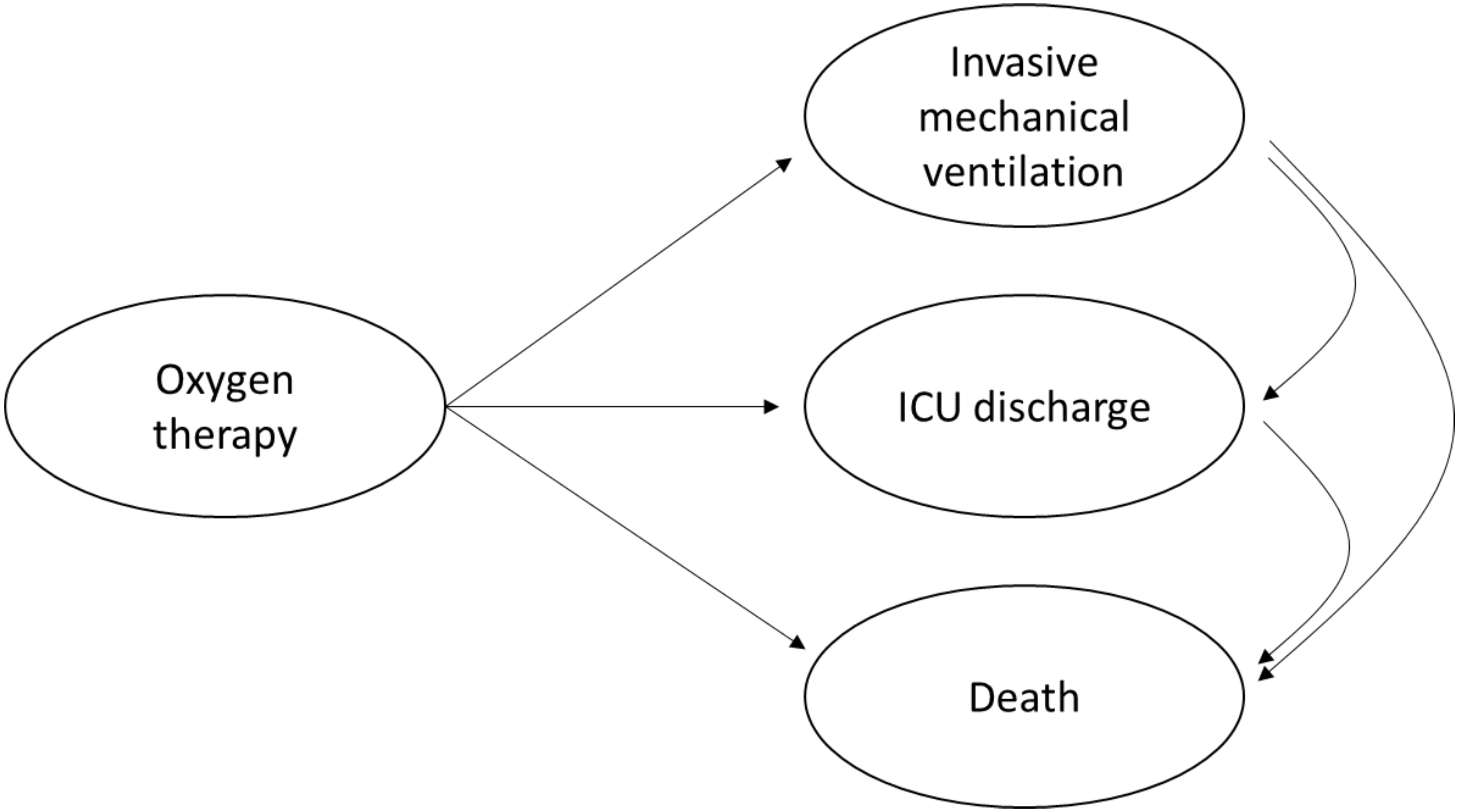

Caption: This figure shows the multistate diagram and possible transitions for the multistate model. Note that patients do not transition from invasive ventilation back to oxygen therapy, so that the “invasive mechanical ventilation” state is more accurately described as the “invasive mechanical ventilation at least once during this ICU admission” state. Similarly, patients cannot be readmitted to ICU, such that the ICU discharge state is better described as “discharged from ICU at least once.”

### Figure S2: Example trace plot from Bayesian multistate model fit

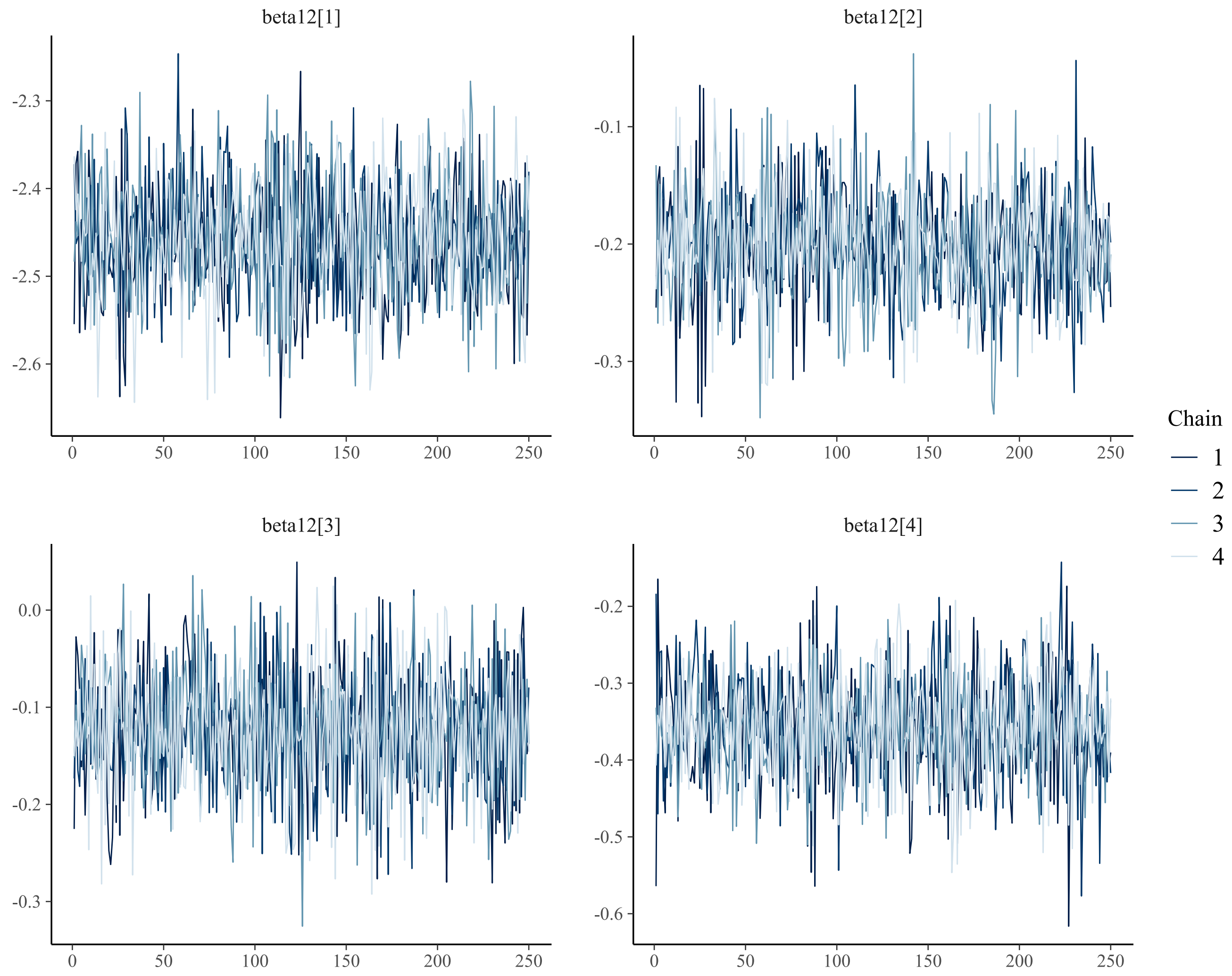

Caption: this plot shows the trace of the posterior samples for the intercept (beta12[1]) and race/ethnicity (beta12[2-4]) parameters from the transition between oxygen therapy and invasive ventilation. Trace plots for other parameters and transitions look similar, with stationary chains exhibiting good mixing and minimal autocorrelation.

#### Figure S3: Correlation plot for posterior distributions of model parameters

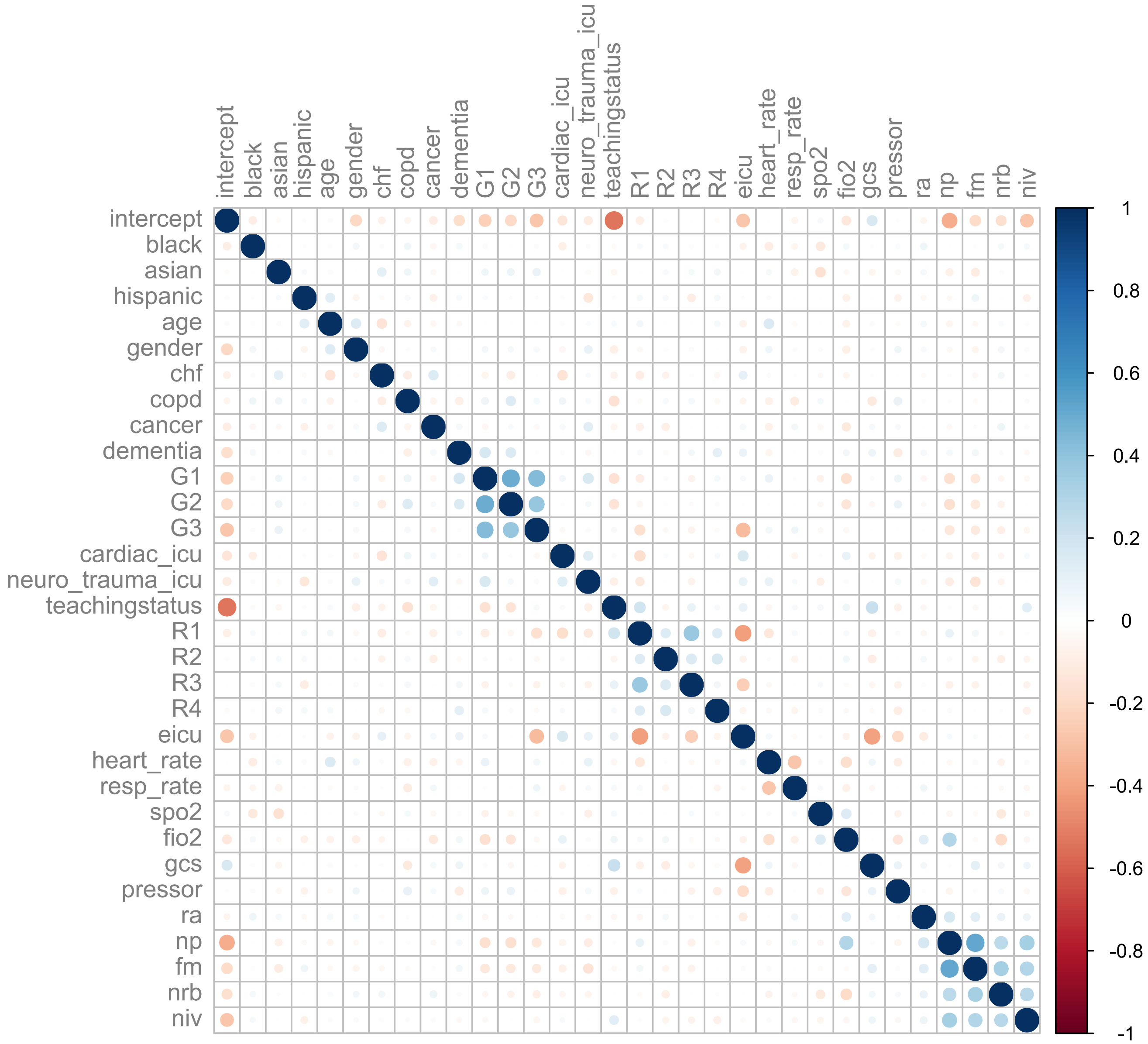

Caption: This plot shows the correlation between posterior parameter values. We include this plot because the models used many predictors, and it was possible that some of the parameters were tightly correlated, in which case their specific parameter values would be less informative. The plot shows that most of the parameters are uncorrelated, and in particular the three race/ethnicity parameters have no meaningful correlation with any of the non-race-ethnicity parameters.

### Figure S4: Forest plot for transition from oxygen therapy to invasive ventilation

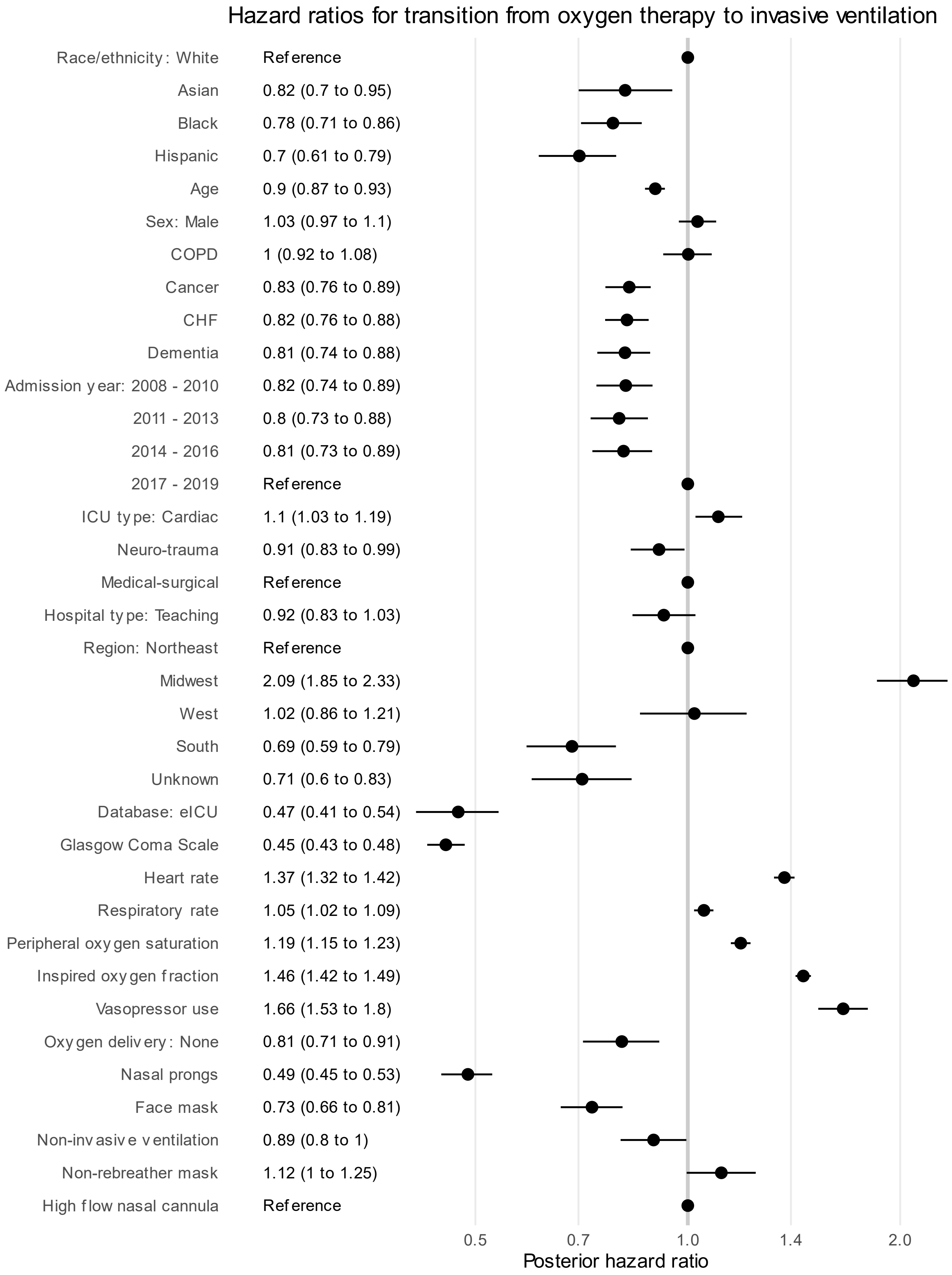

### Figure S5: Forest plot for transition from oxygen therapy to ICU discharge

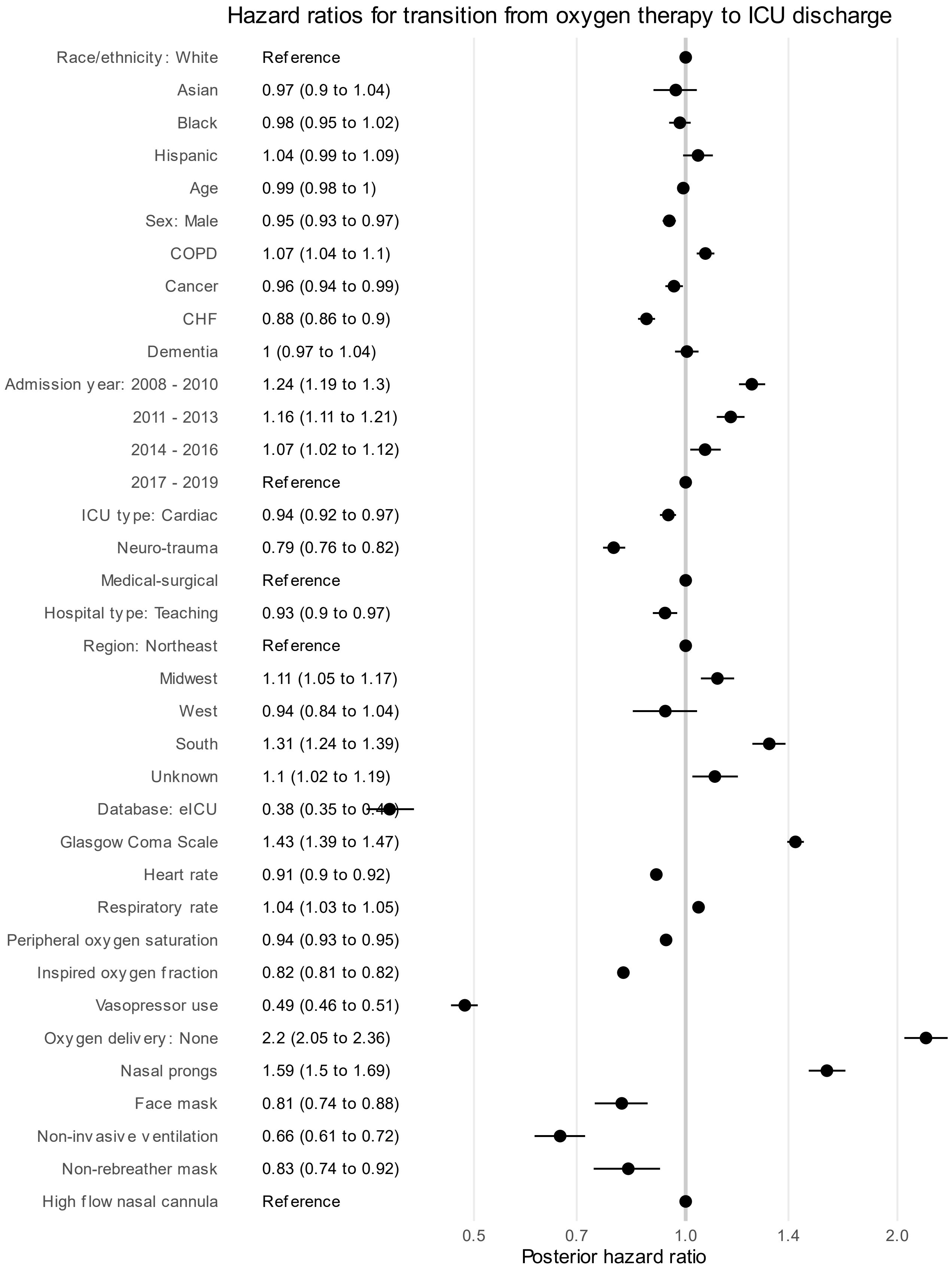

### Figure S6: Forest plot for transition from oxygen therapy to death

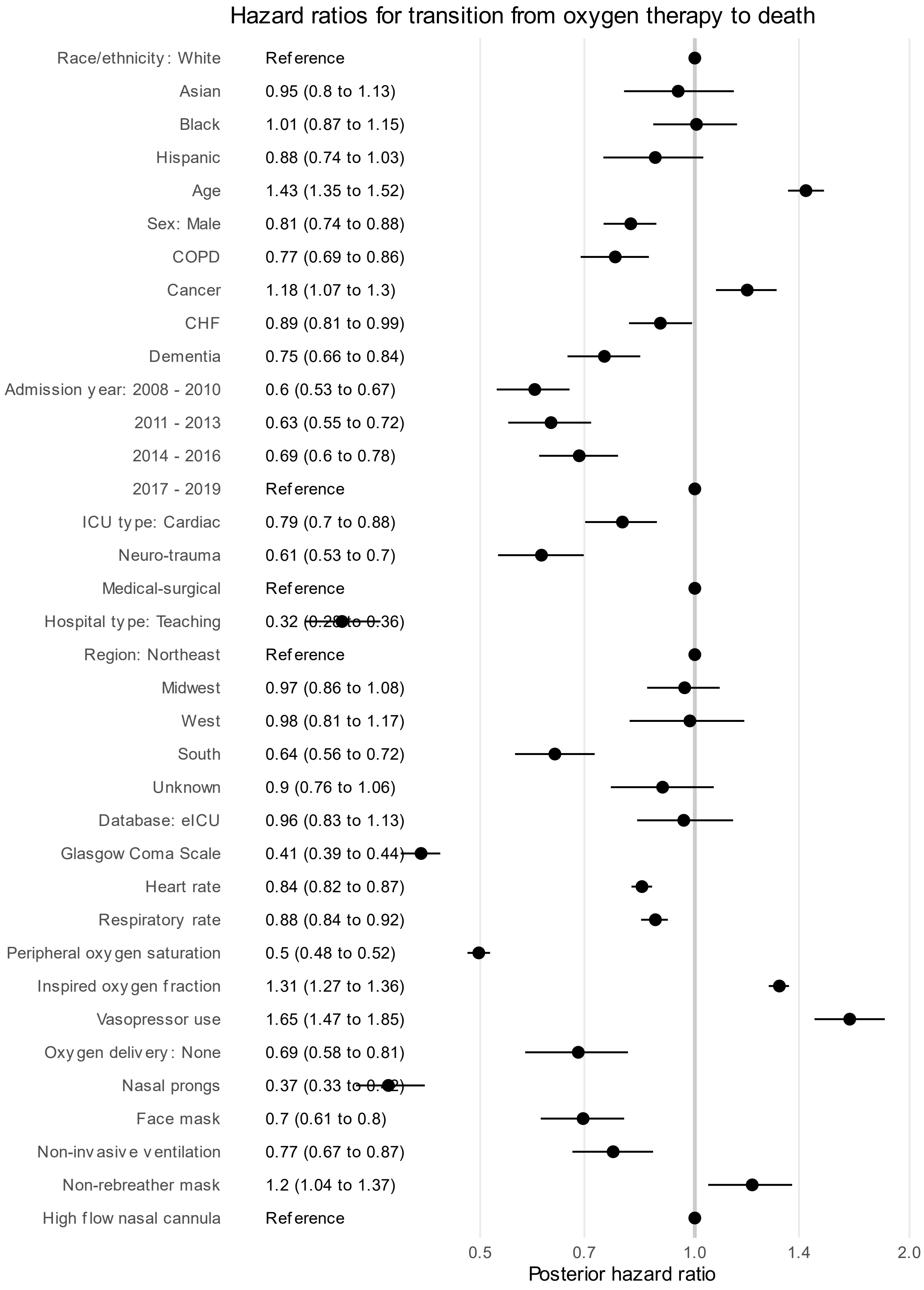

### Figure S7: Forest plot for transition from invasive ventilation to ICU discharge

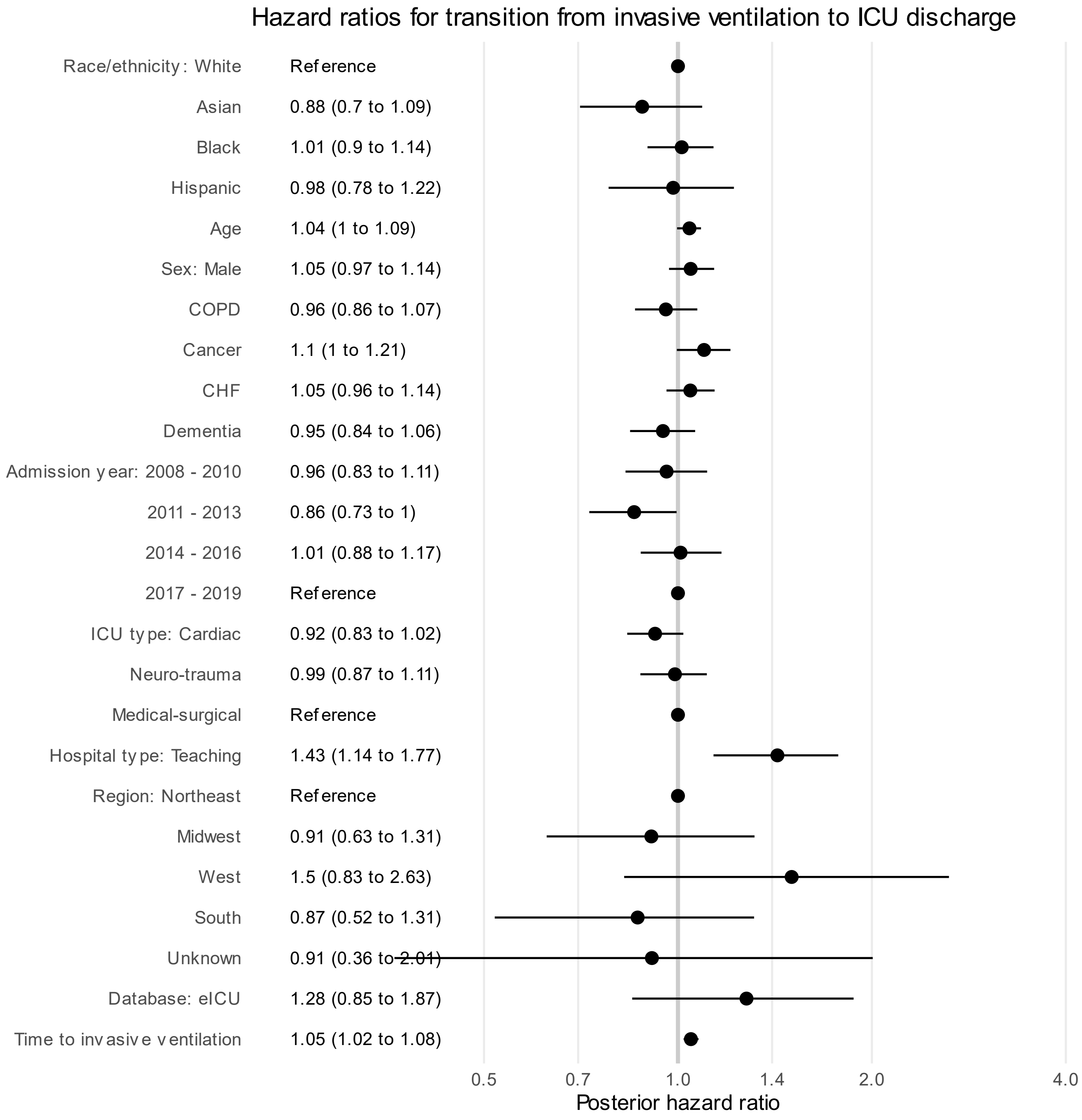

#### Figure S8: Forest plot for transition from invasive ventilation to death

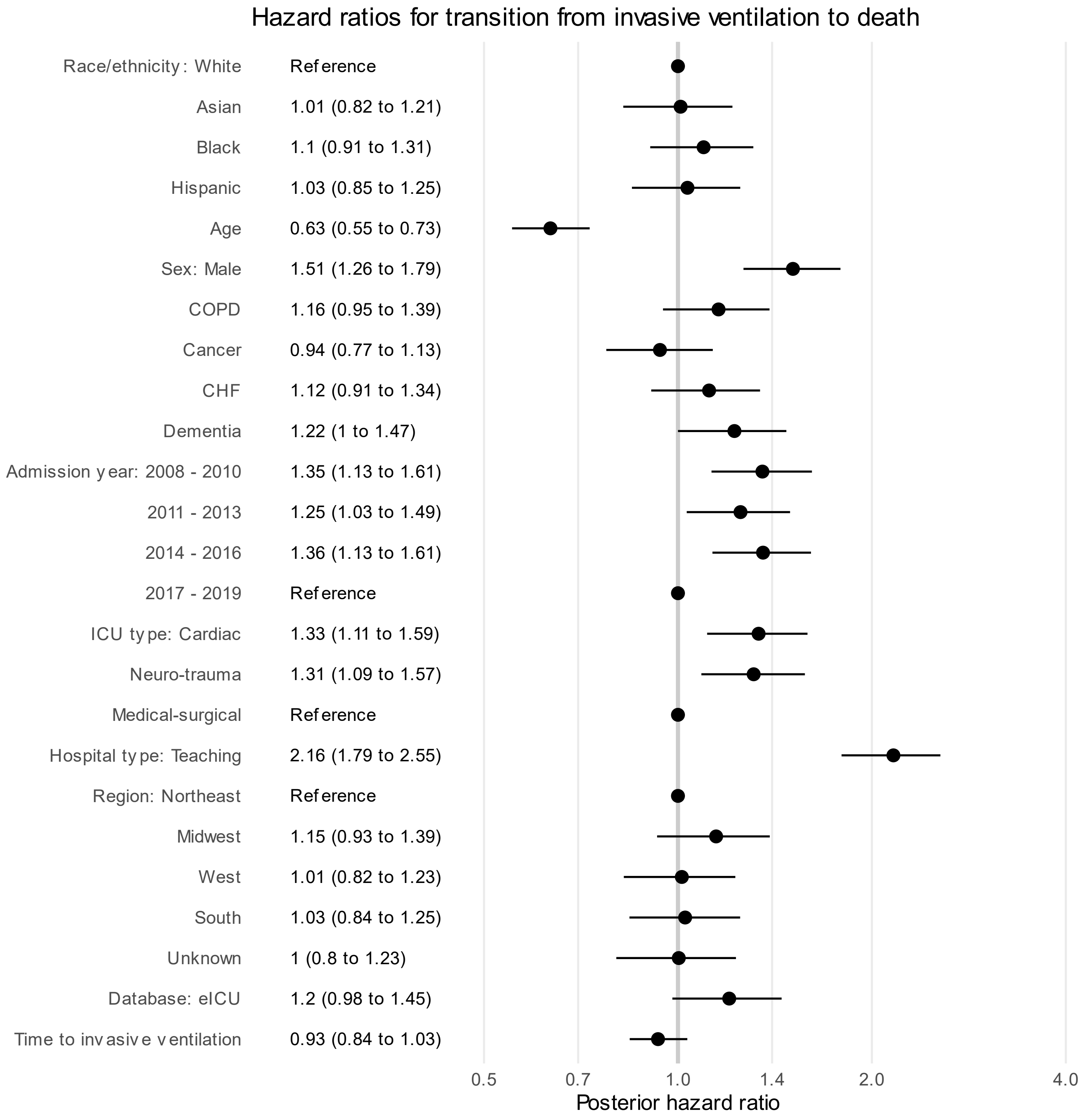

#### Figure S9: Forest plot for transition from ICU discharge to death

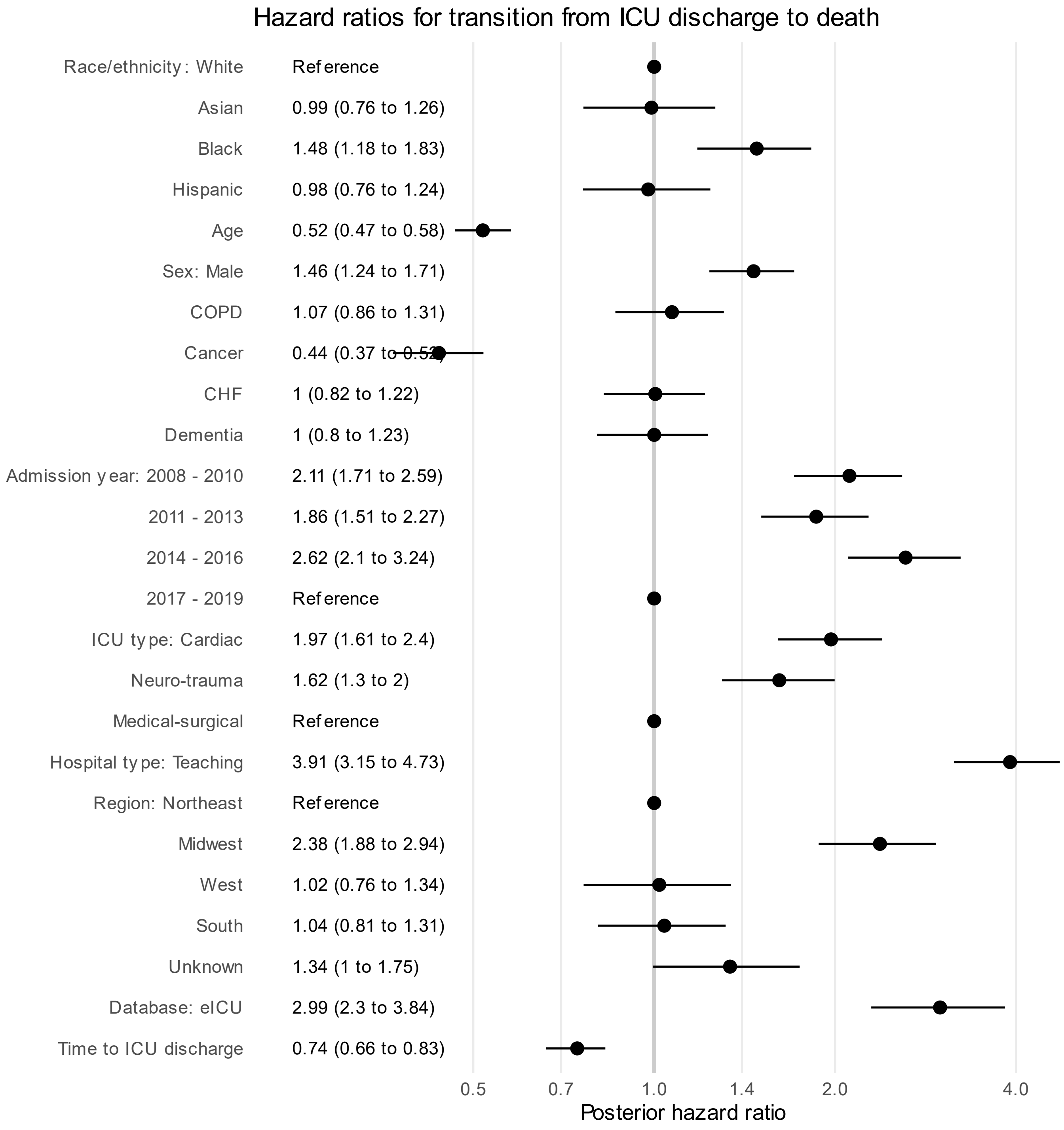

### Figure S10: Forest plot for MIMIC-IV cohort, oxygen therapy to invasive ventilation

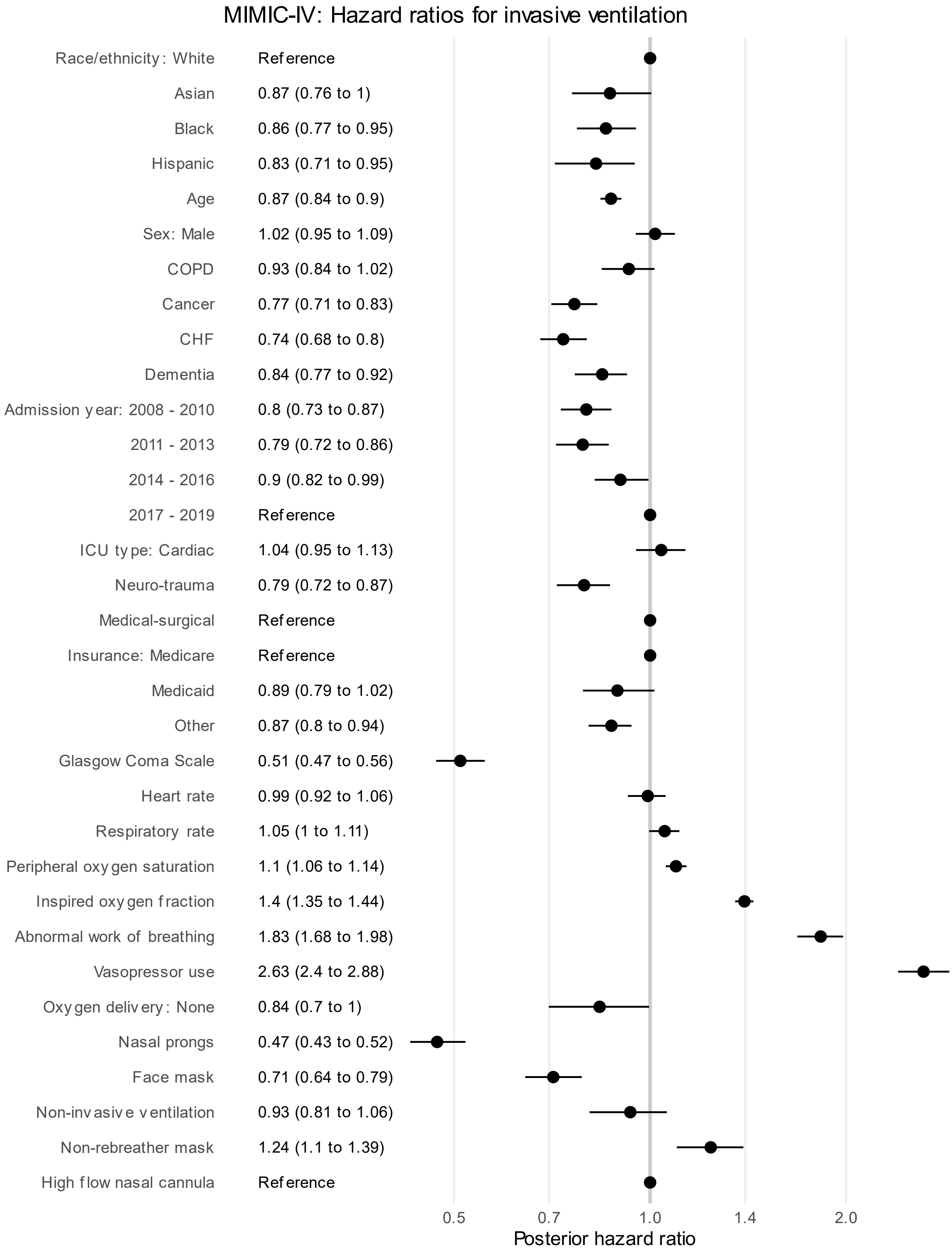

### Figure S11: Forest plot for eICU cohort with same covariates

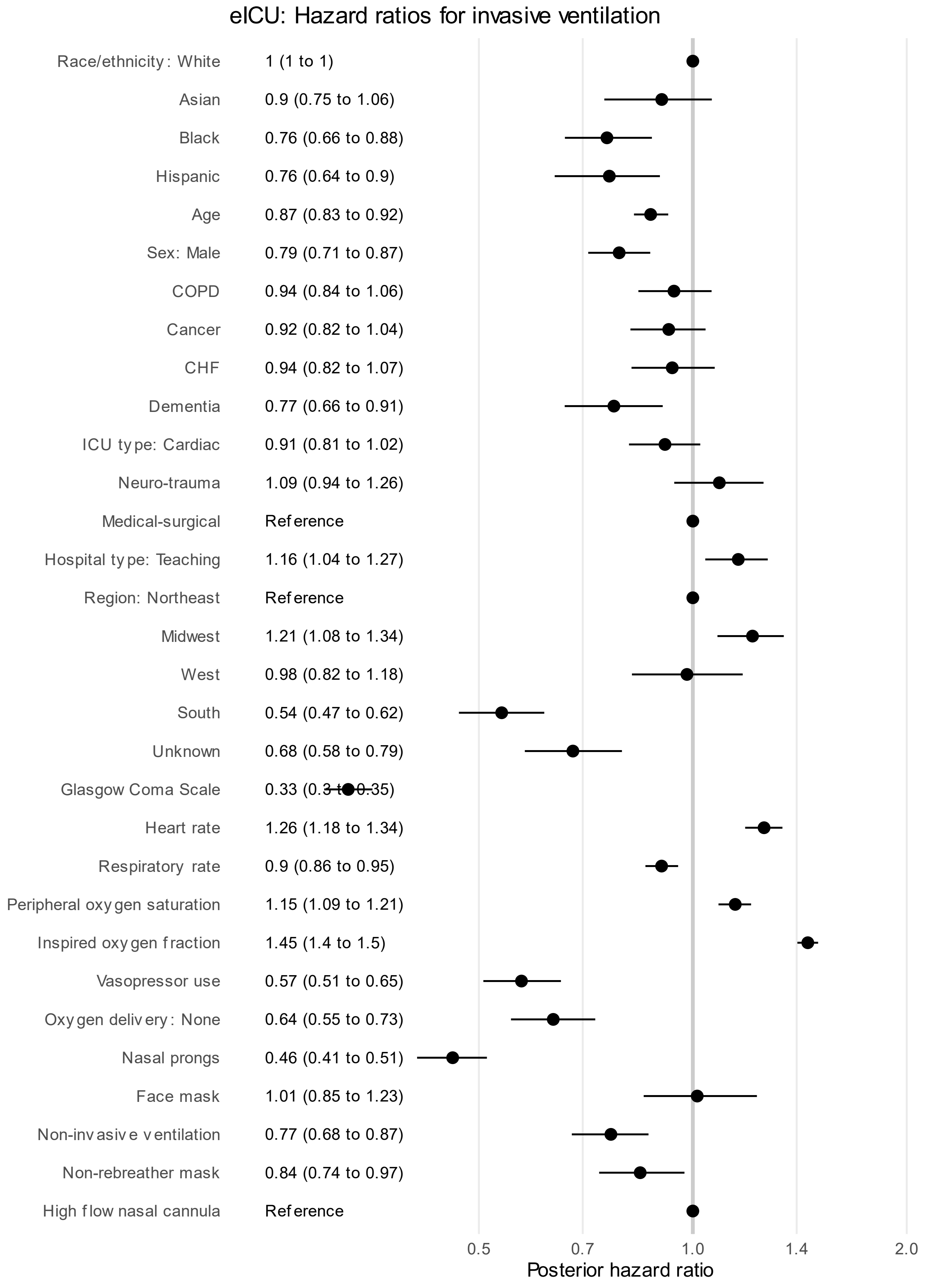

#### Figure S12: Forest plot for eICU cohort with hospital-level covariate

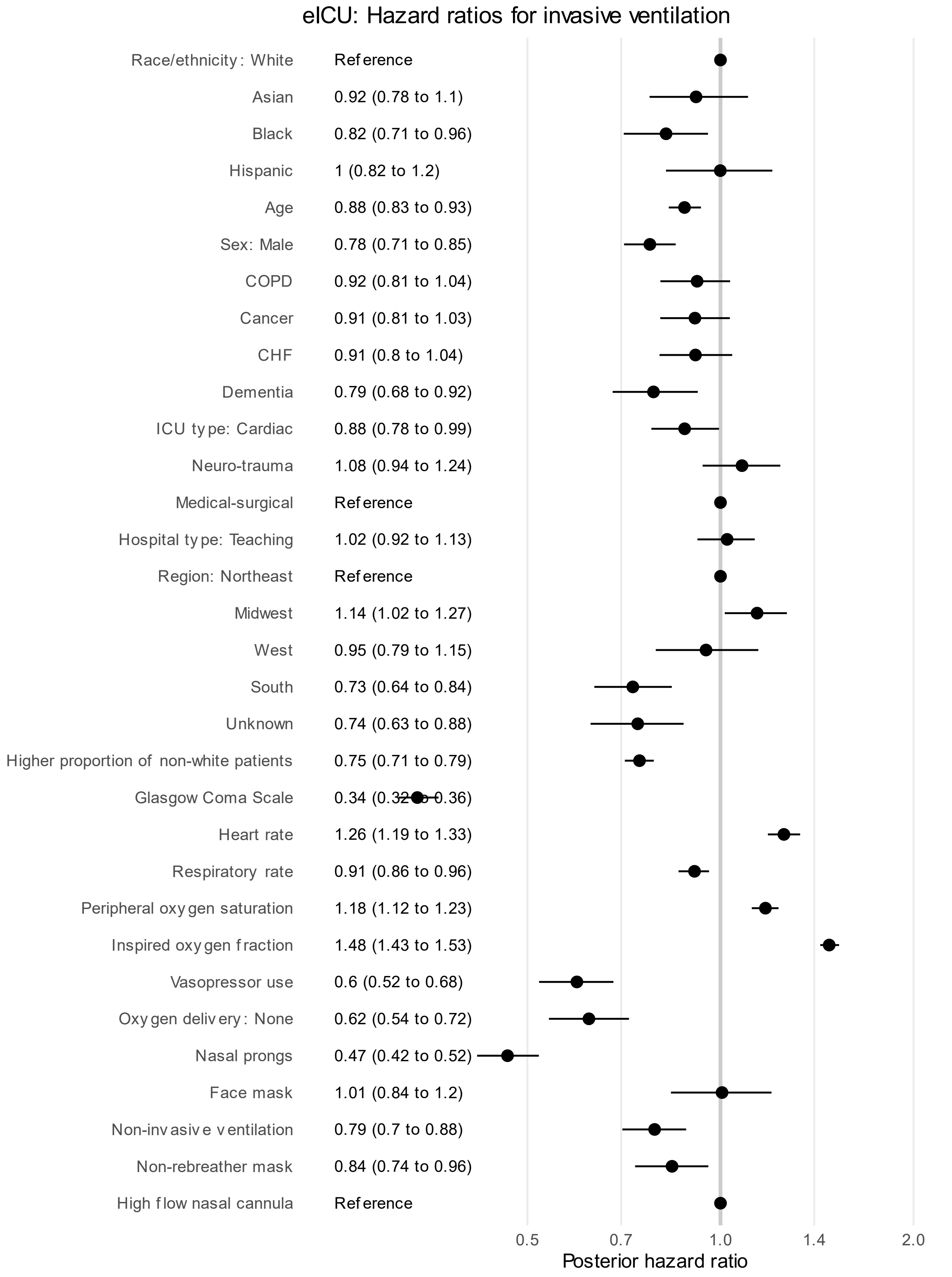
